# Prevention of cyclical resurgences of COVID-19-like pandemics in the long term: What are the trade-offs?

**DOI:** 10.1101/2023.04.16.23288641

**Authors:** Ichiro Nakamoto

## Abstract

Vaccines have facilitated the substantial reduction and containment of COVID-19 transmission in many countries by early 2023. However, the long-term interconnection between vaccines, traits of the pathogen, vaccination strategies, and cases averted/trade-offs of health outcomes is not well understood. Utilizing a compartment-calibrated model, I estimated the aversion/trade-offs effect on six major disease burdens (i.e., total/symptomatic/asymptomatic/hospitalized/ICU/death cases averted) over time conditional on a variety level of scenarios. The findings implied that low-risk immunity profiles of booster doses increased the peak cases averted versus medium- and high-risk counterparts. The effect was most salient for the former paired with enhancing the rollout rate of doses, followed by the medium- and then high-risk scenarios. Positive and temporarily durable aversion effects for the low-risk, in contrast, negative trade-offs and decreasing aversion effects for the suboptimal scenarios were observed. While there are heterogeneities in vaccines, public strategies, social efforts, and other considerations, this work can provide an evidence-based rationale for the long-term trade-off analysis of vaccination.

## Introduction

As of April 2023, a total of over 13 billion doses of COVID-19 vaccines have been administered globally (1-4), which has facilitated the mitigation and containment of SARS-COV-2 transmission by this time. Different countries enacted varied campaigns of vaccination in compliance with the objectives of health policies to balance the trade-offs between the health output of the populations and economic activities (2,3). Retrospectively, vaccines, transmission traits of the pathogen, and vaccination strategies had a combined impact on the risks of infections and subsequent health outcomes (5-10). The data by WHO indicated that there were more than 100 vaccine candidates in clinical usage worldwide by 2023 (1-4). It was of difficulty to exactly capture each of the traits of alternative vaccines in model analyses (11-15). There were heterogeneities and uncertainties regarding the landscape of vaccination, immunity-strength and immunity-waning profiles of doses, and the social effort (1-5, 16-20). Is there a long-term difference in curbing infections and other disease burdens by enacting the booster vaccination versus a primary strategy conditional on the traits of vaccines? How the effect is impacted by the vaccination strategy such as the rollout rate of doses? Furthermore, how to evaluate the potential trade-offs should the immunity profiles of vaccines present heterogeneities? What lessons can be learned from the COVID-19 pandemic to assess the next risk of cyclical resurgence of a novel infectious disease? Should early booster doses for a novel pandemic unveil suboptimal or multi-staged efficacy, where is the potential threshold of trade-offs? Here, I utilized a compartment-calibrated model to mostly capture the risks of infections and reinfections, vaccine inefficacy, immunity strength/waning of vaccines, rate of rollout, and other confounding factors to project the cases averted and trade-offs in the risk appraisal of cyclical resurgences of a COVID-19-like pandemic for six stratified health metrics (i.e., total/symptomatic/asymptomatic/hospitalized/ICU/death cases averted) over a timescale of up to 5 years.

## Methods

Concisely, the transmission of COVID-19-like infectious diseases was divided into multiple compartments, of which primary infections (i.e., initial infections in full susceptibility), reinfections due to waning immunity after recovery (i.e., secondary infections), infections due to inefficacy or multi-staged traits of vaccines after booster vaccination (the rate of which was defined as VI hereafter), immunity waning to susceptibility after booster vaccination (likewise, the rate of which was defined as VS), infections due to immunity waning after booster vaccination (the rate of which was defined as SI), recovery from infections, immunity waning to susceptibility after recovery, rates of vaccination, and vaccination were the major appraised factors in this study (fig. S1, 20-28). The metrics VI, SI, and VS reflected the immunity-strength and immunity-waning profile of booster doses. Theoretically, the greater values of the measures, the lower efficacy of the vaccines (20, 29-33).

The findings in (7) summarized the breakdown of primary infections, including cases requiring hospitalization, hospitalized cases requiring intensive critical care service (ICU hereafter), and death cases for different age groups. Another survey targeting reinfections showed the statistics of death, hospitalizations, and unclear/asymptomatic infections (8). The total cases in this study were comprised of four sections including primary infections, secondary infections, infections due to the inefficacy of vaccines after vaccination, and infections due to immunity waning of vaccines after vaccination. On the other hand, the total cases averted reflected the difference in the number of infections between a primary and a booster strategy, the gap for which could be attributable to multiple factors including the immunity profiles of vaccines, landscape of vaccination, traits of the pathogen, and rate of rollout. To capture the stratified effect of disease burdens in detail, the model divided the initial total cases averted into five stratified health metrics including symptomatic, asymptomatic, hospitalized, ICU, and death cases averted, and the statistics of these measures were calculated respectively based on the statistics data in (7) and (8). Additionally, the model assumed: (a) the population was homogenous and identical population size of each age group; (b) individuals with at least one dose vaccinated were infected with no symptoms (i.e., asymptomatic) due to partial immunity protection of vaccines; (c) the relative infectiousness of other types of infections was identical to the primary infections; (d) the primary vaccination was initiated at week 44, nearly eleven months after the establishment of the transmission; (e) to reflect seasonal variation in transmission, the model based seasonal reproduction numbers in this work on those used in (5), which calibrated values to yield the basic reproduction number of 2.3; (f) no constraint of population size was present;(g) the primary vaccination consisted of two doses, and the booster vaccination consisted of one dose or above.

For purpose of clarity and brevity, the main analysis illustrated the aversion/trade-off effect for a setting with a one-billion population size over an observation period of four years (i.e., equivalently 208 weeks assuming one year consists of 52 weeks) by using a discrete measure of SI from 0.1 to 0.9 in 0.1 increments. I also reflected other varying sizes of populations (i.e., 8e7 to 1.2e9), diverse lengths of observation time (i.e., 160 to 260 weeks), varying SI measured in a continuous scale, and other key parameters in the online interactive dashboard (34). The sensitivity tests on other sizes of populations and intervals of observations were comparable and generalizable. The robustness tests using a series of key parameters implied that the outcomes were consistent for up to 5 years (Supplementary Material and online interactive dashboard).

## Results

### Cyclical trajectory changed with vaccine profiles and vaccination strategies

The model implied that, for a basic reproduction number *R*_0_ = 2.3, the dynamic trajectory of the cyclical pandemic changed with the SI of booster doses over four years. The appraised scenarios could be classified into three categories: high-(Fig. 1, A to C), medium-(Fig. 1, D to F), and low-risk resurgence respectively (Fig. 1, G to I). The risk of cyclical resurgence of the epidemic decreased with the reduction of SI values (Fig. 1, A to F). The lower-risk scenario had fewer times of reoccurrence and the pandemic was contained at an earlier time versus the other two scenarios. On the other, the peak sizes of subsequent resurgence after week 156 (i.e., nearly 80 weeks after the initiation timing of booster doses) for the lower-risk scenario were smaller in magnitude versus the higher-risk counterpart as well (Fig. 1, G, H, and I versus D, E, and F versus A, B, and C).

**Fig. 1.**
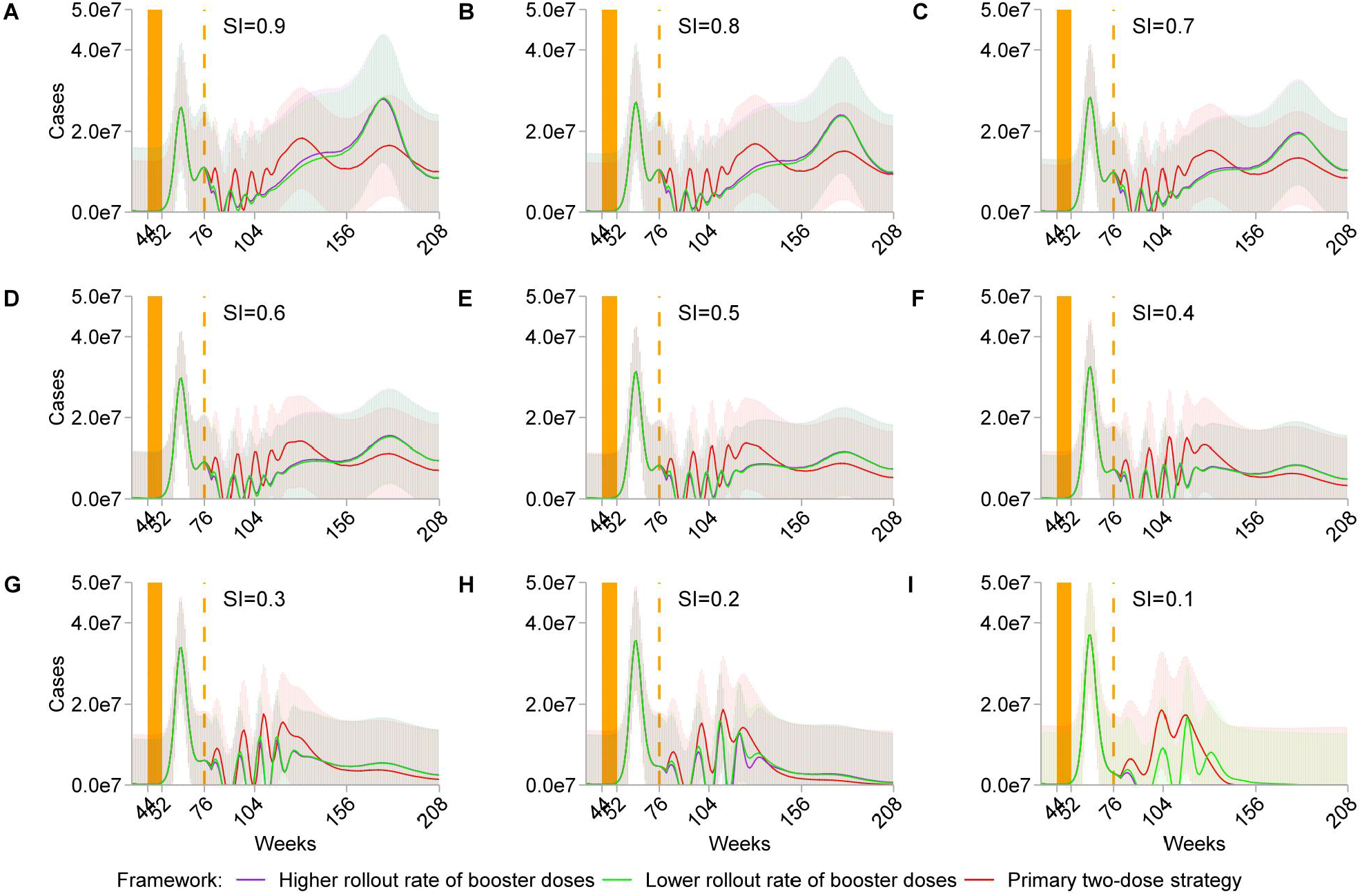
Simulated trajectory of cyclical resurgence. The impact of immunity profiles of vaccines and social efforts on the cyclical occurrences of a COVID-19-like pandemic over four years assuming one year is comprised of 52 weeks. SI varies from 0.9 (A) to 0.1 (I) in 0.1 discrete increments. For each SI scenario, the primary strategy consists of two doses, and the higher and lower rollout rate of booster doses is equal to 4 and 0.5 per year, mimicking scenarios of productive and non-productive distribution respectively. Shaded light-color areas delineate the 95% confidence intervals of the simulations. Orange color bars represent the spacing between the primary doses, and the dashed orange line describes the initiation timing of booster doses. For clarity, I illustrate the trajectory for an 8-week primary spacing and a 24-week booster spacing.

When the immunity strength of vaccines was productive (Fig. 1, I), both primary and booster vaccination contained the pandemic in three years. Distribution of booster doses reduced the peak sizes of the pandemic versus the primary strategy, which was curtailed more in terms of magnitude with the increase in the rollout rate of booster doses. A higher-rate rollout contained the pandemic at an earlier time versus a lower-rate rollout. Asymptotically, the difference in the containment timing of the pandemic between the higher-rate and the lower-rate scenario was 68 weeks (i.e., week 88 versus week 156). When SI increased to 0.2 and the effectiveness of vaccines reduced (Fig. 1, H), the curtailing effect resulting from booster vaccination decreased. No differentiated aversion effect of peak sizes was observed by enhancing the rollout rate of doses versus the lower-rate scenarios. A similar trend was captured when SI was equal to 0.3 (Fig. 1, G). In the medium- and high-risk scenarios, booster vaccination reduced the transmission in the short run (i.e., before week 104), however, cyclical resurgences of the pandemic were likely to occur in the long run (Fig. 1, A to F).

Substantial weekly cases were averted in a short period in the cases where the rollout of booster doses was productive (Fig. 2, I), which facilitated the expeditious mitigation and depletion of the transmission. And the higher-rate scenario was expected to attain a greater aversion in magnitude versus the lower-rate counterpart. For instance, 514026 weekly cases were averted for the former (i.e., a rate equal to 4 per year) versus 333809 weekly cases averted for the latter (i.e., a rate equal to 0.5 per year) at week 80 (Fig. 2, I). When SI increased, the weekly difference of cases averted between the higher- and lower-rate strategy decreased. From a long-term perspective, the positive effect of aversion resulting from booster doses transformed into a negative effect with the increase of SI values (Figs. 2, A to H).

**Fig. 2.**
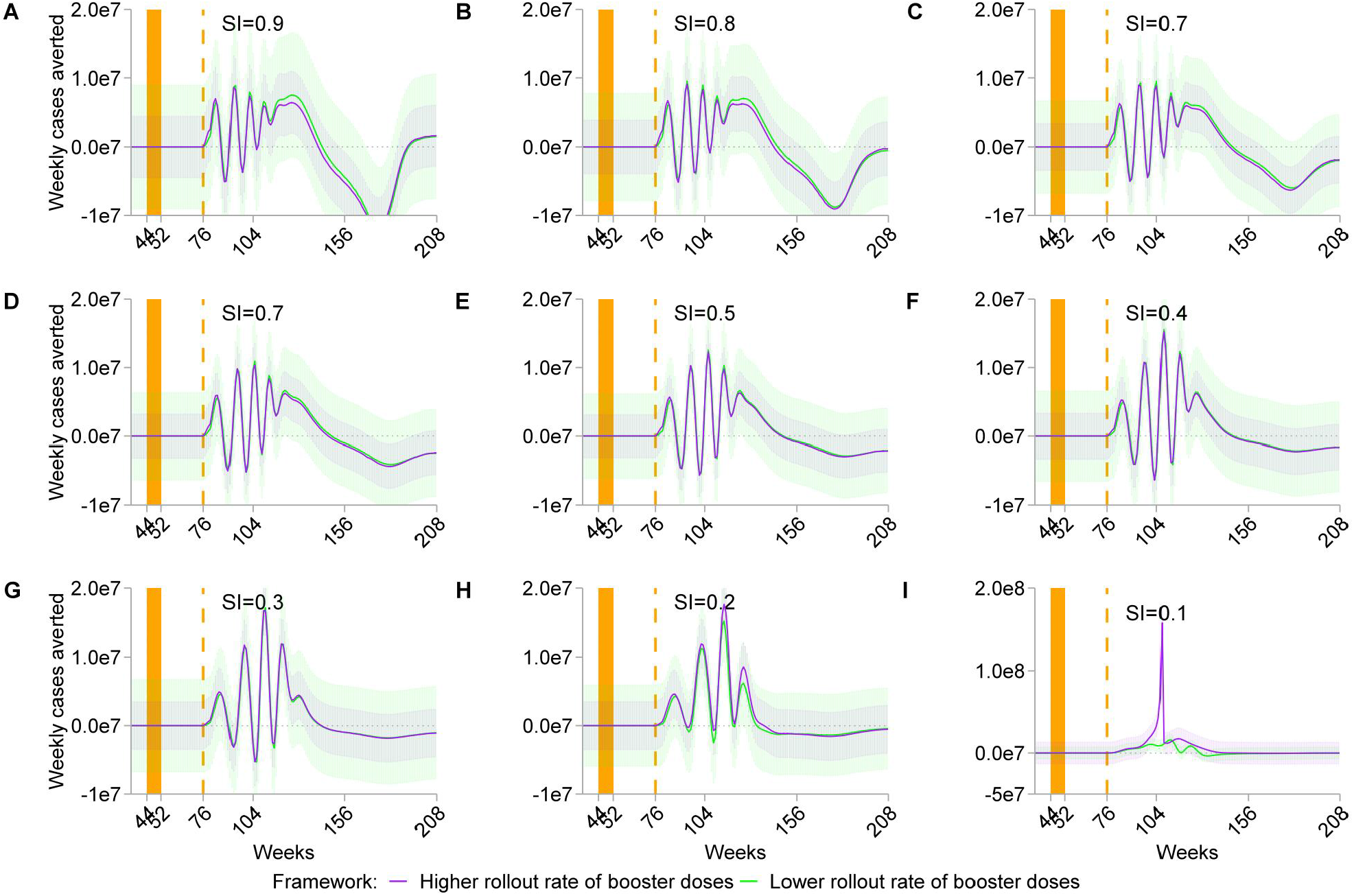
Simulation of weekly cases averted. Weekly new cases averted for SI ranging from 0.9 (A) to 0.1 (I) in 0.1 increments in a four-year timescale assuming one year consists of 52 weeks. For each SI scenario, higher (purple curves and shaded areas) and lower (green curves and shaded areas) rollout rate of booster doses is equal to 4 and 0.5 per year respectively. Shaded light-color areas delineate the 95% confidence intervals of simulations. Orange color bars (weeks 44 and 52) represent the spacing of the primary doses, and dashed orange line (week 76) represents the initiation timing of booster doses.

This section focused on the estimates of the cases averted for the stratified health outcomes including total cases, symptomatic cases, asymptomatic cases, hospitalized cases, ICU cases, and death cases averted over four years. Generally, the results could be divided into three groups including high-(Fig. 3, I1 to I6), medium-(Fig. 3, H1 to H6), and low-salient scenarios (Fig. 3, A1 to G6) conditional on the magnitude of aversion and the difference between productive and non-productive rollout of doses respectively.

**Fig. 3.**
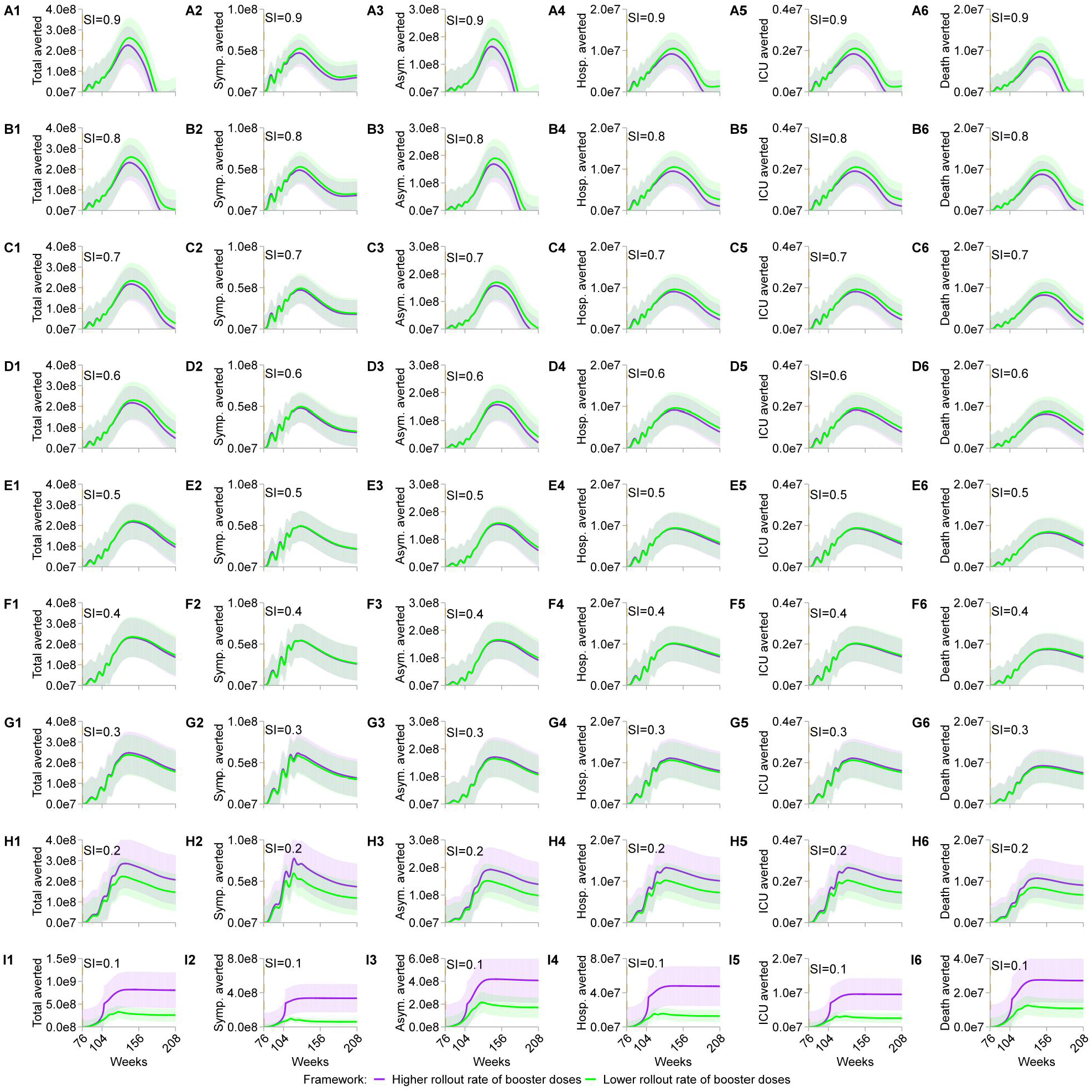
Simulation stratified cases averted for six major disease burdens. Simulation of the aversion effect for major metrics of health outcomes with SI ranging from 0.9 (A1 to A6) to 0.1 (I1 to I6) in 0.1 increments. All scenarios sketch the cumulative cases averted over four years. “Total averted” scenario denotes the total cases averted; “Symp. averted” depicts the symptomatic cases averted; “Asym. averted” represents the asymptomatic cases averted; “Hosp. averted” represents the hospitalization cases averted; “ICU averted” outlines the number averted of patients needing intensive care unit; “Death averted” delineates the death cases averted. For each SI scenario, higher (purple curves and shaded areas) and lower (green curves and shaded areas) rollout rates of booster doses are equal to 4 and 0.5 per year respectively. Shaded light colors sketch the 95% confidence intervals of the simulations. A more detailed continuous measure of SI, differentiated rollout rates of booster doses, diverse booster spacing, varying length of observation time, and other key parameters are delineated in the online interactive dashboard.

In the high-salient cases (Fig. 3, I1 to I6), each of the disease burdens yielded a non-decreasing and salient aversion in terms of magnitude accrued over time. For instance, the curves of the total cases averted observed the peak date (i.e., the timing of yielding the maximum cases averted) of week 145 for a higher-versus earlier week 128 for a lower-rate vaccination. The former was expected to yield a cumulative saturation of 8.190465e+08 (95% CI: 434346410, 1203746552, hereafter CI) cases averted, and the latter 3.282357e+08 (CI: 194672962, 461798377) cases averted at the peak points respectively (Fig. 3, I1, and Supplementary data file S1). The aversion rate was roughly 81.9% for the higher- and 32.8% for the lower-rate rollout respectively divided by an assumed one-billion population size taking into account the risks of reinfections, the difference in vaccination strategies, the traits of disease transmission, and the traits of vaccines. For each stratified metric under the same level of SI, the cases averted remained at the commensurate level relative to the peak sizes at the end of appraisal time, implying a robust and consistent aversion effect when SI values were lower. In a salient-effect scenario, a higher-capacity rollout could capture an effect of more than double-fold versus the lower-capacity counterpart (Fig. 3, I1 to I6, and Supplementary data file S1). When SI transferred to the medium-salient scenario in which SI=0.2 (Fig. 3, H1), the cases averted for the total cases dropped versus the high-salient scenario in which SI=0.1(Fig. 3, I1). Asymptotically, the peak date shifted to week 137 for the higher- and week 133 for the lower-rate scenario, the peak size of which was 285826987(CI: 167652930, 404001045) for the former and 223255075 (CI: 134128570, 312381581) for the latter respectively (Fig. 3, H1, and Supplementary data file S1). This implied a nearly 28.6% and 22.3% aversion rate under the same size of the population previously. In the medium- and high-salient cases, the size averted for the total cases decreased over time after the peak-date point (Fig. 3, A1, B1, C1, D1, E1, F1, G1, and Supplementary data file S1). In a similar vein, at SI=0.3, the peak date observed was at the commensurable point of time for the higher-versus for the lower-rate scenario, and 247459708 (CI: 146289736, 348629681) versus 2.378337e+08 (CI: 140641248, 335026083) in magnitude respectively (Fig. 3, G1, and Supplementary data file S1). It could be seen that with the increase of SI, the gap of cases averted between productive and non-productive rollout lessened or reversed at certain points (Fig. 3, A1, B1, C1, D1, E1, F1, and Supplementary data file S1), suggesting the limited effect of rollout capacity enhancing and the complexity of the transmission when vaccines were ineffective. The greater values of SI, the faster reduction in the cases averted over time after the peak points, indicating the earlier terminating of the positive effect (Fig. A1, B1, C1, D1, E1, F1, G1, H1, and Supplementary data file S1).

Consistently, the trending observed for total cases averted applied to the other five stratified disease burdens including symptomatic, asymptomatic, hospitalized, ICU, and death cases averted when SI=0.1 (Fig. 3, A2 to I6). For instance, the estimated peak date for maximum symptomatic cases averted was week 146 for the higher-versus week 115 for the lower-rate scenario, and the peak size averted was 3.373556e+08(CI: 175563367, 499147914) versus 9.617363e+07(CI: 62891532, 129455724) (Fig. 3, I2, and Supplementary data file S1), implying a 33.74% and 9.62% aversion rate respectively. In a comparable analysis, asymptomatic cases averted observed the peak date of week 146 versus week 128, and the peak size averted of 4.189621e+08 (CI: 224273867, 613650426) versus 2.154589e+08 (CI: 127649753, 303268108) respectively (Fig. 3, I3, and Supplementary data file S1), indicating a 41.9% and 21.55% aversion rate. As for hospitalization cases averted, the data yielded the peak date of week 146 versus week 128, and the peak size averted of 4.795871e+07 (CI: 25191367, 70726047) versus 1.562443e+07 (CI: 9089754, 22159110) (Fig. 3, I4, and Supplementary data file S1), the aversion rates of which were 5.00% and 1.56% respectively. The peak date for ICU cases averted was estimated at week 146 versus week 128, and the peak size averted captured a magnitude difference of 9591741 (CI: 5038273, 14145209) versus 3.124887e+06 (CI: 1817950, 4431822) (Fig. 3, I5, and Supplementary data file S1). Analogous calculation yielded aversion rates of 1% and 0.3%. Finally, death cases averted estimated the saturation points of week 146 versus 129, and the saturation size averted of 2.745773e+07 (CI: 14578065, 40337389) versus 1.249529e+07 (CI: 7122512, 17868064) (Fig. 3, I6, and Supplementary data file S1), for which the aversion rates were 2.75% and 1.25%. To sum up, when SI was low, enhancing the rollout rate of booster doses increased the cases averted and the effect of aversion was robust over time after the peak dates for symptomatic, asymptomatic, hospitalized, ICU, and death cases. The peak dates were reached approximately after 61 to 70 weeks of the rollout for the higher-rate scenario and 39 to 52 weeks for the lower-rate scenario, suggesting the complexity of the disease transmission.

In the scenarios where the SI value increased to 0.2, the cases averted for each of the stratified disease burdens reduced in magnitude (Fig. 3, H1 to H6). Representatively, symptomatic cases averted observed the homogeneous peak date of week 119, whereas the peak size of 77612795 (CI: 49790759, 105434831) for the high-versus 59550066 (CI: 39163409, 79936723) for the low-capacity scenario (Fig. 3, H2, and Supplementary data file S1), which resulted in a 7.76% and 5.96% aversion rate respectively. Likewise, the simulation forecast the peak date of week 140 versus week 136 for asymptomatic cases averted, and the peak size averted of 192469959 (CI: 112526792, 272413128) versus 1.516442e+08 (CI: 90813414, 212474984) (Fig. 3, H3, and Supplementary data file S1), producing a 19.25% and 15.16% aversion rate. For the hospitalization scenario, the projected values were week 144 versus week 131, and 12883256 (CI: 7331233, 18435280) versus 10212040 (CI: 6039796, 14384285) (Fig. 3, H4, and Supplementary data file S1), yielding the aversion rate of 1.29% and 1%. The ICU scenario observed week 132 versus week 131, and peak size averted of 2654109 (CI: 1543704, 3764514) versus 2042408 (CI: 1207959, 2876857) (Fig. 3 H5, and Supplementary data file S1), from which computing the aversion rate of 0.3% versus 0.2%. Finally, the peak date of week 142 versus week 139 for death cases averted, and the peak size averted of 10742189 (CI: 6086897, 15397481) versus 8460058 (CI: 4877025, 12043092) (Fig. 3, H6, and Supplementary data file S1), representing the aversion rate of 1% and 0.8%. The peak dates were reached with a slightly earlier time for the higher-rate scenario versus SI=0.1.

When the SI value increased to 0.3 or greater, the cases averted decreased further for each stratified disease burden (Fig. 3, A1 to G6, and Supplementary data file S1). The gap of cases averted for each metric reduced or reversed between higher- and lower-capacity scenarios when SI inflated. Taking the scenario SI=0.3 as an illustration, the peak size averted for symptomatic cases curtailed to 61559292 (CI: 38861942, 84256642) versus 58618482 (CI: 37067324, 80169641) (Fig. 3, G2, and Supplementary data file S1). The asymptomatic cases averted observed similar trending, the peak size averted downsized to 1.703002e+08 (CI: 100268604, 240331865) versus 1.641417e+08 (CI: 96645891, 231591011) (Fig. 3, G3, and Supplementary data file S1). This lowered to 1.106625e+07 (CI: 6444994, 15687515) versus 1.059409e+07 (CI: 6169283, 15018895) for hospitalized cases (Fig. 3, G4, and Supplementary data file S1). And the peak ICU cases averted shrank to 2213250 (CI: 1288998, 3137502) versus 2.118818e+06 (CI: 1233856, 3003778) (Fig. 3, G5, and Supplementary data file S1). Finally, peak death cases averted dropped to 9249231 (CI: 5263545, 13234918) versus 8.902579e+06 (CI: 5064754, 12740403) respectively (Fig. 3, G6, and Supplementary data file S1). The positive role played by enhancing the rollout rate of booster doses reduced with the increase of SI values. Greater values of SI also curtailed the magnitude of aversion and terminate the positive aversion effect earlier versus lower values.

### Trade-offs when immunity profiles of vaccines transformed to suboptimal

I estimated the trade-offs for the stratified cases averted over four years when the immunity profiles of booster doses changed to suboptimal by varying SI and VI from 0 to 1.0 using 50 simulations for each measure using the previous population size ceteris paribus (Fig. 4, A to F). The main analysis illustrated the outcomes for the higher-rate rollout scenario, and the lower-rate scenario implied a similar effect of trending (Fig. S25, A to F). The parameters used in the simulation were outlined in the Supplementary Material (Table S2). Consistently, all the stratified health metrics including total, symptomatic, asymptomatic, hospitalized, ICU, and death cases averted observed positive trade-offs when the values of SI and VI were lower (i.e., the areas where values asymptotically lower than 0.2), and negative trade-offs when suboptimal (i.e., the areas where values asymptotically greater than 0.2).For instance, when VI=0.3 and SI=0.3 ceteris paribus, the estimate asymptotically yielded the negative trade-offs of -5e8 cases for total cases averted (Fig. 4, A), -4e7 cases for symptomatic cases averted (Fig. 4, B), -4e8 cases for asymptomatic cases averted (Fig. 4, C), -2e7 cases for hospitalized cases averted (Fig. 4, D), -5e6 cases for ICU cases averted (Fig. 4, E), and -2e7 cases for death cases averted using a population size of 1e9 respectively (Fig. 4, F). These values reflected the rates of social cost of 50%, 4%, 40%, 2%, 0.5%, and 2% respectively. The greater values of SI and VI, the larger effect of negative trade-offs.

**Fig. 4.**
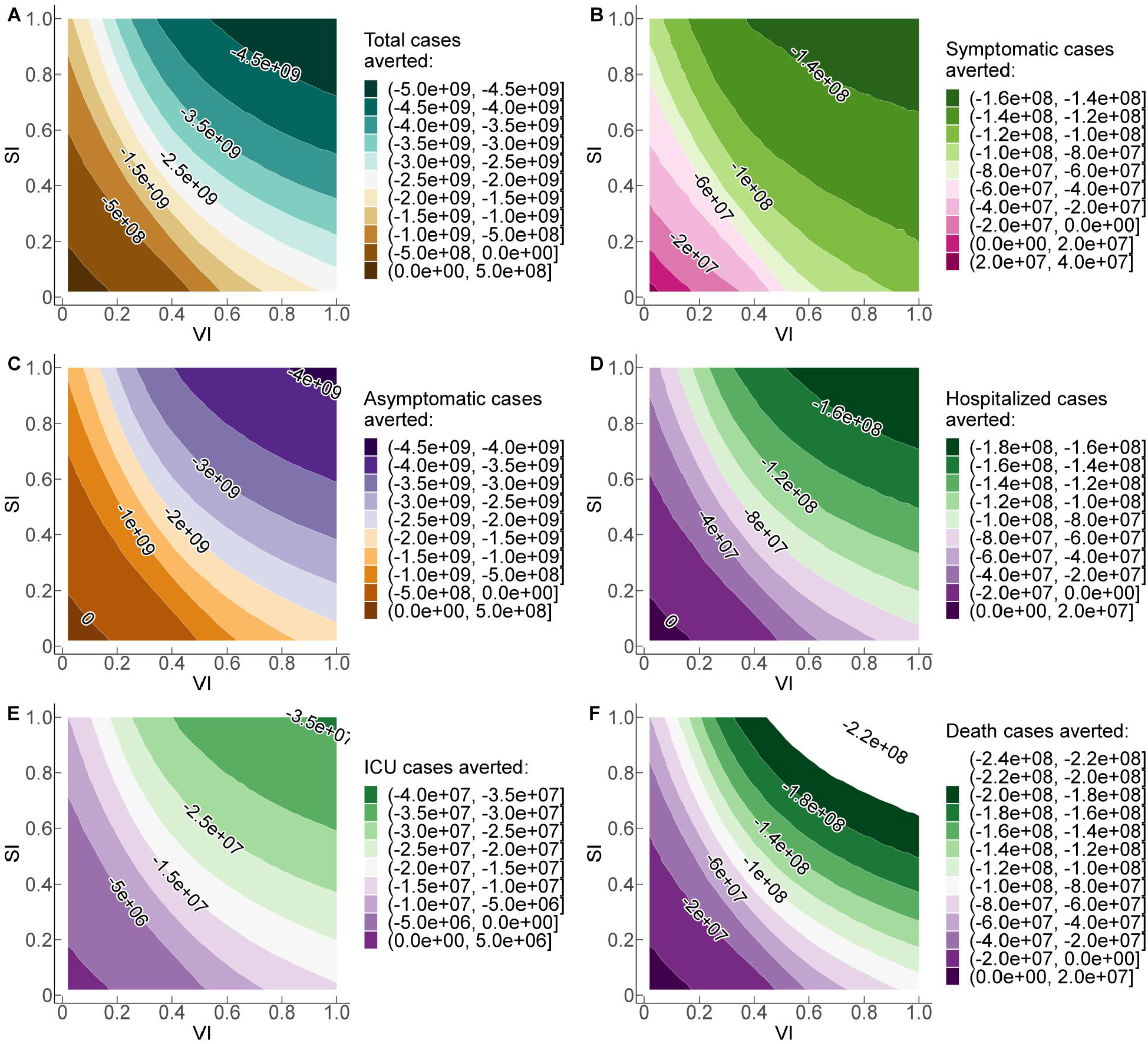
Simulation trade-offs of disease burdens varying SI and VI from 0 to 1. Simulation of the cumulative stratified cases averted over four years with SI and VI ranging from 0 to 1.0 using 50 simulations for each metric respectively. The population size is equal to 1e9. Positive numbers on each curve illustrate the positive cases averted for each health outcome, and negative numbers depict the negative trade-off when VI and SI transform to sub-optimal ceteris paribus. The rollout rate of booster doses is 4 per year, representing the productive rollout scenario.

In this section, I performed analysis on the trade-offs for the same stratified health metrics with SI and VS varying from 0 to 1.0 using 50 simulations for each respectively (Fig. 5, A to F). Consistent with previous findings, all measures observed a positive aversion effect when VS and SI were lower. The effect transformed to negative trade-offs when VS and SI were suboptimal ceteris paribus. For instance, when VS=0.3 and SI=0.6, the range of trade-offs was approximately [-1e9,-5e8] for total cases averted (Fig. 5, A), [-6e7,-4e7] for symptomatic cases averted (Fig. 5, B), [-1e9,-5e8] for asymptomatic cases averted (Fig. 5, C), [-4e7, -2e7] for hospitalized cases averted (Fig. 5, D), [-1e7, -5e6] for ICU cases averted (Fig. 5, E), and [-4e7, -2e7] for death cases averted respectively (Fig. 5, F). These reflected the rate ranges of social cost of [50%, 100%], [4%, 6%], [50%, 100%], [2%, 4%], [0.5%, 1%], and [2%, 4%] respectively. The parameters used in the simulation and the outcomes were summarized in the Supplementary Material (Table S3, Fig. S26, A to F). For constant values of SI in the higher range, increasing the values of VS raises the magnitude of negative trade-offs, suggesting the salient role that VS played in the transmission of diseases.

**Fig. 5.**
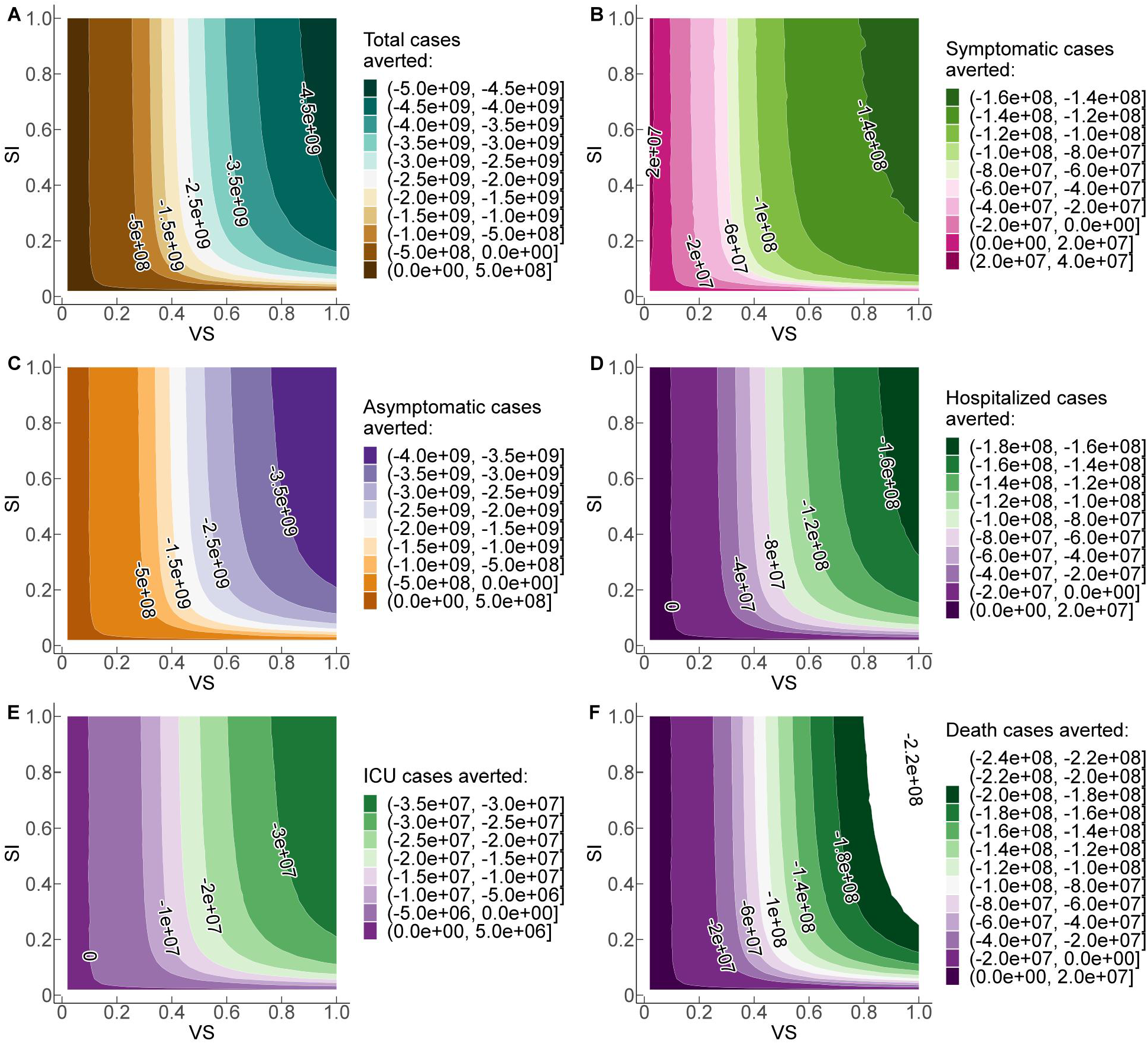
Simulation trade-offs of disease burdens with varying immunity profiles using VS and SI. Simulation of the cumulative stratified cases averted over four years with SI and VS ranging from 0 to 1.0 using 50 simulations for each metric respectively. The population size is equal to one billion. Positive numbers on each curve illustrate the positive cases averted for each health burden, and negative numbers depict the negative trade-off when VI and SI are sub-optimal ceteris paribus. The rollout rate of booster doses is 4 per year, representing the productive rollout scenario.

### Positive cases averted when immunity profiles of vaccines are productive

Next, I estimated how the values of SI and VI contributed to the positive effect of aversion by concentrating on the interval [0, 0.1] where the immunity profiles of vaccines are productive, for both measures (Fig. 6, A to F). The results implied that lower values of VI and SI reduced the social cost and yielded a greater magnitude of effect versus higher-value counterparts for total, symptomatic, asymptomatic, hospitalized, ICU, and death cases averted. For instance, when VI=0.05 and SI=0.05, the cases averted were asymptotically 8e7 for total cases (Fig. 6, A), 1e7 for symptomatic cases (Fig. 6, B), 6e7 for asymptomatic cases (Fig. 6, C), 3e6 for hospitalized cases (Fig. 6, D), 6e5 for ICU cases (Fig. 6, E), and 3e6 for death cases respectively (Fig. 6, F). These produce the aversion rate of 8%, 1%, 6%, 0.3%, 0.06%, and 0.3% respectively.

**Fig. 6.**
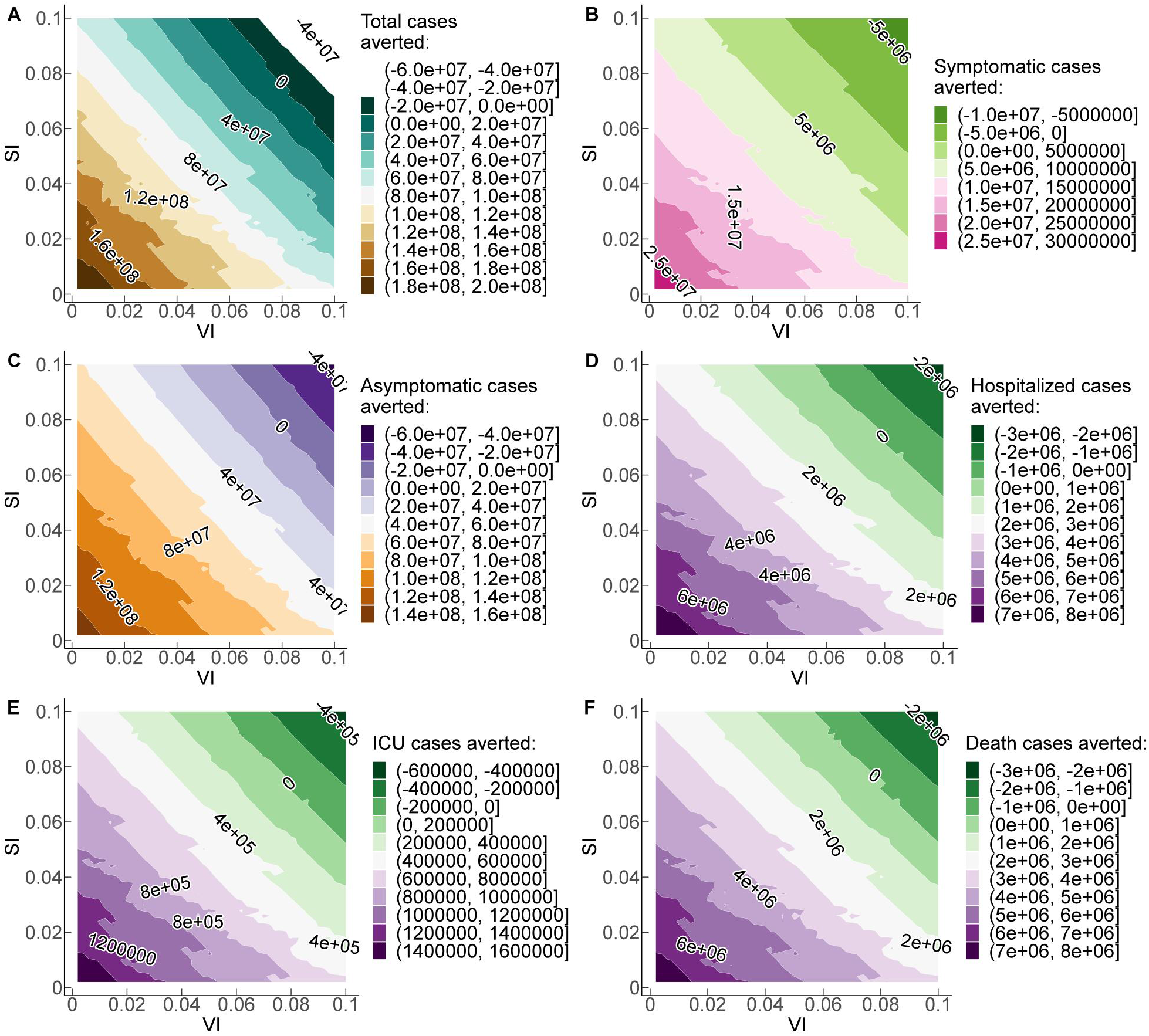
Simulation stratified cases averted for disease burdens with SI and VI between 0 and 0.1. Simulation of the stratified cases averted over four years for the six major health burdens with SI and VI varying from 0 to 0.1 using 50 simulations for each metric respectively. The population size is equal to 1e9. Positive numbers on each curve illustrate the positive aversion effect, and negative numbers depict the negative trade-off for each health outcome when VI and SI are sub-optimal ceteris paribus. The rollout rate of booster doses is 4 per year, representing the productive rollout scenario.

The estimates of SI and VS varying from 0 to 0.1 were analyzed in a similar vein (Fig. 7, A to F). For a constant value of SI=0.04 and population size of 1e9, calibration of VS from 0.07 to 0.05 asymptotically engendered positive cases averted of 2e7 for total cases (Fig. 7, A), 2e6 for symptomatic cases (Fig. 7, B), 2e7 for asymptomatic cases (Fig. 7, C), 1e6 for hospitalized cases (Fig. 7, D), 2e5 for ICU cases (Fig. 7, E), and 1e6 for death cases (Fig. 7, F). In contrast, when VS=0.05 and SI=0.05, the cases averted were asymptotically [6e7,8e7] for total cases (Fig. 7, A), [8e6,1e7] for symptomatic cases (Fig. 7, B), [4e7, 6e7] for asymptomatic cases (Fig. 7, C), [2e6, 3e6] for hospitalized cases (Fig. 7, D), [4e5, 6e5] for ICU cases (Fig. 7, E), and [2e6, 3e6] for death cases respectively (Fig. 7, F). These outcomes reflected the rates of aversion effect of [6%, 8%], [0.8, 1%]. [4%, 7%], [0.2%, 0.3%],[0.04%, 0.06%], and [0.2%, 0.3%] respectively.

**Fig. 7.**
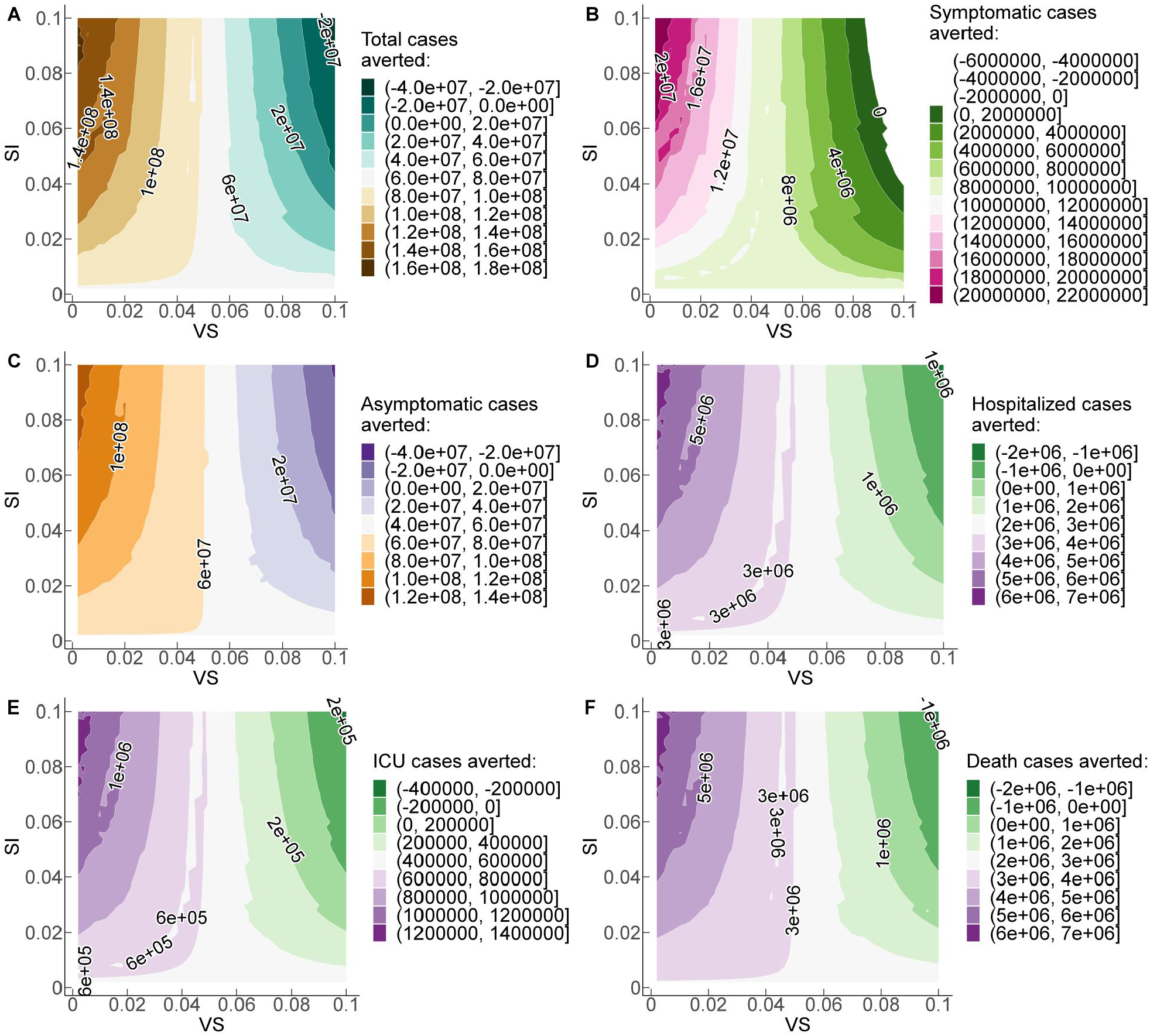
Simulation stratified cases averted for major disease burdens with SI and VS. Simulation of the stratified cases averted over four years for SI and VS varying from 0 to 0.1 using 50 simulations for each metric respectively. The population size is equal to 1e9. Positive numbers on each curve illustrate the positive aversion effect, and negative numbers depict the negative trade-off for each health outcome when VS and SI are sub-optimal ceteris paribus. The rollout rate of booster doses is 4 per year, representing the productive rollout scenario.

### Sensitivity tests of cases averted using major parameters

#### One-way sensitivity analysis

I performed a series of sensitivity analyses on the stratified cases averted for all six disease burdens by employing key parameters of the transmission, traits of the pathogen, traits of vaccines, and vaccination strategies. For each of the parameters, changing a specific metric ceteris paribus to project the update of aversion trajectory.

##### Rate of waning to susceptibility after vaccination

Experiments conducted on recovered patients found heterogeneous levels of detectable SARS-CoV-2 neutralizing antibodies, implying a differentiated rate of immunity waning to susceptibility after vaccination (7-16, 25-29). I tested the sensitivity across high-, medium-, and low-risk scenarios for primary doses, and the major findings were robust (figs. S2 to S7).

##### Rate of birth and death

Considering the years of evaluation, the population might change during the period (20, 22-28). I estimated two scenarios corresponding to a slower rate and a faster rate of population change respectively. The outcomes did not change qualitatively (figs. S8 to S9).

##### Vaccination rate of primary doses

Studies in (20-26,29-34) suggest that the vaccination rate of doses could impact the trajectory of disease transmission. I tested these scenarios and the outcomes remained qualitatively the same as in the main analysis (figs. S10 to S11).

##### Vaccination rate of booster doses

To test how the change in the vaccination rate of booster doses played roles in the trajectory of aversion effect (35-42), a greater rate and a slower rate were used respectively (fig. S12 to S13). Other rollout rates of booster doses can be found in the online interactive dashboard (34). The robustness test for this parameter also supported the consistency.

##### Timing of vaccination of booster doses

The timing of the vaccination potentially exerted an impact on the transmission and hence the aversion track (20, 43-48). I tested two scenarios by shifting the timing of booster doses to an earlier/delayed point respectively, and the results were qualitatively unchanged (figs. S14 to S15). As expected, delaying the rollout of booster doses yielded a curtailed aversion effect.

##### Initial size of infections

I tested the scenarios with greater and smaller sizes of initial infections respectively (20, 49-54), and the outcome remained at the commensurate level (figs. S16 to S17). The tests using a different initial ratio of secondary over primary infections also supported this robustness (34).

##### Size of population

The model did not place constraints on the population size. The main analysis presented the outcomes corresponding to a setting with one-billion population size (20,35-38). I also tested the scenario where the population size was equal to one-tenth of the one-billion size and the analysis implied a comparable outcome (fig. S18 to S22). For the effects on other sizes of populations, please refer to the online interactive dashboard (34).

##### Rate of waning to secondary susceptibility after recovery

Patients either obtained natural and/or vaccination immunity after recovery, which reduced over time and individuals could be exposed to reinfections at a later time (20, 25-33). I tested the rate of waning to secondary susceptibility after recovery and the results qualitatively identified the robustness(34).

### Multiway sensitivity analysis

Additionally, I selected seven parameters (susceptibility rate to infection after immunity of dose 1&2 wanes, rate of waning to susceptibility after dose 1&2 vaccination, rate of infection after dose 1&2 vaccination, rate of waning to secondary susceptibility after recovery) for which the uncertainties were of concerns and tested two combinations of these major parameters (figs. S23 to S24). The cases averted were quite robust in these analyses as well (figs. S23 to S24). For more detailed information regarding the length of observation time, continuous measure of SI, sizes of populations, initiation timing of booster vaccination, other key parameters, and how the responsiveness of the results to the parameters, please refer to the online interactive dashboard (34).

## Discussion

The findings suggest that, assuming *R*_0_ = 2.3, booster vaccines with low-risk immunity profiles could produce a salient effect for major disease burdens including total, symptomatic, asymptomatic, hospitalized, ICU, and death cases averted up to 5 years, the magnitude of which is followed by medium- and then high-risk counterparts respectively. Enhancing the rollout rate of booster doses yields a greater and more durable aversion effect over time after the peak points versus lower-rate rollout scenarios when SI was low, and the effect was reduced with the increase of SI values. In a setting with a one-billion of population size and SI=0.1, the peak aversion rate of total cases was approximately 81.9% for the higher-rate rollout (i.e., a rate equal to 4 per year) versus 32.8% for the lower-rate rollout (i.e., a rate equal to 0.5 per year), 33.74% versus 9.62% for peak symptomatic cases, 41.9% versus 21.55% for asymptomatic cases, 5.00% versus1.56% for hospitalization cases, 1% versus 0.3% for ICU cases, 2.75% versus 1.25% for death cases respectively. These rates reduced to 28.6% versus 22.3%, 7.76% versus 5.96%, 19.25% versus 15.16%, 1.29% versus 1%, 0.3% versus 0.2%, 1% and 0.8% respectively when SI=0.2 and lower when SI values were greater. Enhanced social effort facilitated the rapid mitigation and containment of the pandemic when SI was low, whereas the gap of the aversion effect was reduced when SI increased (20-23, 39-45). The greater values of SI, the faster reduction in the cases averted over time after the peak points and the earlier termination of the positive aversion effect. When immunity profiles of vaccines (e.g., SI, VI, and VS) transformed from productive to suboptimal, negative trade-offs were observed for all disease burdens. The findings were consistent with other vaccine modeling studies (20-28,54-55). The greater values of SI, VI, and VS, the larger the negative trade-offs generated. The results suggest that enhanced social effort through the rollout improvement of booster doses facilitated the control of disease transmission when the immunity profiles of doses were productive.

Many countries used nearly three years or more to control and/or mitigate the spread of the COVID-19 pandemic, and the diminishing of the transmission reflected substantial collaborations and effort (35,39-45). Here, a compartment-refined model was utilized to determine the cases averted and trade-offs for stratified disease burdens up to 5 years. The outcomes were consistent over time for total, symptomatic, asymptomatic, hospitalized, ICU, and death cases averted. I have assumed a plain framework for capturing the critical traits of SARS-CoV-2 booster vaccines or other infectious diseases that would partially share similar traits with SARS-CoV-2. Nearly 20 years passed since the first outbreak of the SARS pandemic in 2003 (1-4,20-24), and it could be difficult to pinpoint the next risk. However, it is expected that when vaccines are available should a novel infectious disease emerge, more scientific knowledge about the mechanisms, including the viral load trajectory, the relationship with dynamic infectiousness, and the vaccination strategies would accrue over time, allowing refinement of the modeling and projection feasible (20-23,49-53). In practice, the magnitude of the effect can be updated by calibrating the parameters to new settings as more information unfolds.

This study has several limitations. First, the model assumed an equal and constant infectiousness profile for different types of infections including primary, secondary, and other infections. Scientific knowledge of the information for viruses and subsequent variant sub-lineages is incomplete and more observations are necessary, although some studies have made progress in this direction (26-31, 46-51). If the infectiousness profiles of different infections change over time, then the findings need to update when more accurate and dynamic data are available (20-25). Second, the model assumed a constant immunity-waning rate, infection/reinfection rate, and other rates, however, these conditions could modify in practice conditional on the transmission complexity of the pathogen and the traits of vaccines (20,32-39). As more detailed measures are available, the model can be recalibrated to reflect the updated information. Third, the model neither captured geographical/age/contact nor the primary-spacing differences, however, these factors potentially exerted a non-negligible impact on the stratified disease burdens (20-24, 31-35). As the population makeup, geographic traits, and social contacts could be very different across settings, and similarly the primary-dose spacing for vaccines. Potentially these were the key to determining region-, age-, contact-, and spacing-based estimates of cases averted and trade-offs. Fourth, I used the rates of various disease burdens published in (7) and (8), and the seasonal reproduction number published in (20) to project the potential impact, but these statistics might vary vastly over time and/or over locations. In these cases, models need to be refined and calibrated to reflect the temporal and geographic context. And the generalization of the results needed to be exercised with caution. Fifth, I assumed a two-dose primary strategy, which could be different for the differentiated practices in countries. Further, I used three critical traits of vaccines to estimate the stratified effect, whereas other important measures connected with the stratified disease burdens potentially present. For instance, occupation and vaccination hesitancy was potentially linked to an increased risk of asymptomatic and symptomatic infections in certain settings (12-18, 33-37). Furthermore, several studies (38-46) identified that, as a result of healthcare in-equilibrium, individuals from racial and ethnic minority groups were exposed to increased risk versus their counterparts in certain countries. These are critical considerations in further directions of studies to identify more detailed effects for the stratified disease burdens.

Finally, I have explored the simplest framework, which can only provide a general implication of the aversion potential for the stratified measures of health outcomes under different scenarios should a COVID-19-like pandemic occur. Including more complicated evolutionary components (20, 41-48) in the model is an important direction for future work. Heterogeneities of the pathogen, population makeup, vaccines, and vaccination strategies likely change chronically, which may have important impacts on the trajectory of peak dates, peak sizes, and trade-offs (20,35-38). The results can asymptotically provide an estimate of the traits of the pathogen, traits of the transmission, vaccines paired with social effort for stratified metrics of disease burdens, which can be used as long-term evidence-based guidance to the retrospective threshold appraisal of vaccination strategies, social gains/costs and the stratified trade-off analysis in the long term.

## Data Availability

All data needed to estimate the conclusions in the study are present in the paper, the Supplementary Materials, and/or the Supplementary Data. The code used in the analysis is available at https://github.com/wcg305/prevention_cyclical_resurgence_COVID-19. The interactive dashboard addressing the sensitivity tests and key parameters discussed in the manuscripts is accessible from the address https://ichironakamoto.shinyapps.io/analysis_aversion_COVID-19-like_pandemic. Additional data related to this paper may be requested from the corresponding author.

## Acknowledgments

I.N. acknowledges W. Chen for helpful discussions regarding the calibration analysis of parameters.

## Author contributions

I.N. conceived the study, developed the model, wrote the coding and performed analysis, and produced and revised the manuscript. All authors contributed to the final draft.

## Supplementary Materials

Please refer to the supplementary material for detailed information regarding the mathematical model, the parameters used in the simulations, and the results of the sensitivity tests.

