## supplementary file for "Prevention of cyclical resurgences of COVID-19-like pandemics in the long term: What are the trade-offs?"

Ichiro Nakamoto*

**This PDF file includes:**

Figs. S1 to S26
Tables S1 to S3
Supplemental Materials and Methods
References (#1 to #55)

**Other Supplementary Materials for this manuscript include the following:**
Data file S1.

### Mathematical model

This supplementary text outlines the framework of mathematical models, the flowchart of infections, and equilibrium solutions of the transmission dynamics. I herein include the sensitivity test outcomes for the six stratified disease burdens including total, symptomatic, asymptomatic, hospitalized, ICU, and death cases averted respectively. The definitions and values of parameters utilized in the simulations are illustrated in Table S1 to S3. More detailed information regarding key parameters can be found in the online interactive dashboard (34). Suppose a susceptible-vaccinated-infected-recovered-susceptible [SVIRS] model and a population of flexible size N (20-22), in which each individual can be re-infected and the types of infections are multiple (Fig. S1).

The model governing the epidemic transmission dynamics can be expressed as follows:

$$\frac{dS_P}{dt} = \mu - \beta S_P\left[I_P + \alpha I_S + \alpha_V I_V + \alpha_1 I_{S_1} + \alpha_2 I_{S_2} + \alpha_3 I_{S_3}\right] - (s_{vax1}\nu + \mu)S_P \tag{1}$$

$$\frac{dI_P}{dt} = \beta S_P\left[I_P + \alpha I_S + \alpha_V I_V + \alpha_1 I_{S_1} + \alpha_2 I_{S_2} + \alpha_3 I_{S_3}\right] - (\gamma + \mu)I_P \tag{2}$$

$$\frac{dR}{dt} = \gamma\left[I_P + I_S + I_V + I_{S_1} + I_{S_2} + I_{S_3}\right] - (\delta + \mu)R \tag{3}$$

$$\frac{dS_S}{dt} = \delta R - \varepsilon\beta S_S\left[I_P + \alpha I_S + \alpha_V I_V + \alpha_1 I_{S_1} + \alpha_2 I_{S_2} + \alpha_3 I_{S_3}\right] - (s_{vax1}\nu + \mu)S_S \tag{4}$$

$$\frac{dI_S}{dt} = \varepsilon\beta S_S\left[I_P + \alpha I_S + \alpha_V I_V + \alpha_1 I_{S_1} + \alpha_2 I_{S_2} + \alpha_3 I_{S_3}\right] - (\gamma + \mu)I_S \tag{5}$$

$$\frac{dV_1}{dt} = s_{vax1}\nu S_P + c s_{vax1}\nu S_S - \varepsilon_{V_1}\beta V_1\left[I_P + \alpha I_S + \alpha_V I_V + \alpha_1 I_{S_1} + \alpha_2 I_{S_2} + \alpha_3 I_{S_3}\right] - (\omega s_{vax2} + \rho_1 + \mu)V_1 \tag{6}$$

$$\frac{dV_2}{dt} = d s_{vax1}\nu S_S + \omega s_{vax_2} V_1 - \varepsilon_{V_2}\beta V_2\left[I_P + \alpha I_S + \alpha_V I_V + \alpha_1 I_{S_1} + \alpha_2 I_{S_2} + \alpha_3 I_{S_3}\right] - \left(\omega_1 s_{vax_3} + \rho_2 + \mu\right)V_2 \tag{7}$$

$$\frac{dV_3}{dt} = (1 - c - d)s_{vax1}\nu S_S + \omega_1 s_{vax3} V_2 - \varepsilon_{V_3}\beta V_3\left[I_P + \alpha I_S + \alpha_V I_V + \alpha_1 I_{S_1} + \alpha_2 I_{S_2} + \alpha_3 I_{S_3}\right] - (\rho_3 + \mu)V_3 \tag{8}$$

$$\frac{dI_V}{dt} = \beta(\varepsilon_{V_1}V_1 + \varepsilon_{V_2}V_2 + \varepsilon_{V_3}V_3)\left[I_P + \alpha I_S + \alpha_V I_V + \alpha_1 I_{S_1} + \alpha_2 I_{S_2} + \alpha_3 I_{S_3}\right] - (\gamma + \mu)I_V \tag{9}$$

$$\frac{dS_{S_1}}{dt} = \rho_1 V_1 - \varepsilon_1\beta S_{S_1}\left[I_P + \alpha I_S + \alpha_V I_V + \alpha_1 I_{S_1} + \alpha_2 I_{S_2} + \alpha_3 I_{S_3}\right] - \mu S_{S_1} \tag{10}$$

$$\frac{dS_{S_2}}{dt} = \rho_2 V_2 - \varepsilon_2\beta S_{S_2}\left[I_P + \alpha I_S + \alpha_V I_V + \alpha_1 I_{S_1} + \alpha_2 I_{S_2} + \alpha_3 I_{S_3}\right] - \mu S_{S_2} \tag{11}$$

$$\frac{dS_{S_3}}{dt} = \rho_3 V_3 - \varepsilon_3 \beta S_{S_3}\left[I_P + \alpha I_S + \alpha_V I_V + \alpha_1 I_{S_1} + \alpha_2 I_{S_2} + \alpha_3 I_{S_3}\right] - \mu S_{S_3} \qquad (12)$$

$$\frac{dI_{S_1}}{dt} = \varepsilon_1 \beta S_{S_1}\left[I_P + \alpha I_S + \alpha_V I_V + \alpha_1 I_{S_1} + \alpha_2 I_{S_2} + \alpha_3 I_{S_3}\right] - (\gamma + \mu) I_{S_1} \qquad (13)$$

$$\frac{dI_{S_2}}{dt} = \varepsilon_2 \beta S_{S_2}\left[I_P + \alpha I_S + \alpha_V I_V + \alpha_1 I_{S_1} + \alpha_2 I_{S_2} + \alpha_3 I_{S_3}\right] - (\gamma + \mu) I_{S_2} \qquad (14)$$

$$\frac{dI_{S_3}}{dt} = \varepsilon_3 \beta S_{S_3}\left[I_P + \alpha I_S + \alpha_V I_V + \alpha_1 I_{S_1} + \alpha_2 I_{S_2} + \alpha_3 I_{S_3}\right] - (\gamma + \mu) I_{S_3} \qquad (15)$$

To implement vaccination campaigns, the conditions of the following need to be satisfied:

$$S_{vax1} = \begin{cases} 0, & t < t_{vax1} \\ 1, & t \geq t_{vax1} \end{cases} \qquad (16)$$

$$S_{vax2} = \begin{cases} 0, & t < t_{vax2} \\ 1, & t \geq t_{vax2} \end{cases} \qquad (17)$$

$$S_{vax3} = \begin{cases} 0, & t < t_{vax3} \\ 1, & t \geq t_{vax3} \end{cases} \qquad (18)$$

In a disease-free equilibrium, no incidence of infections occurs and thus the system can be rephrased as follows:

$$\frac{dS_P}{dt} = \mu - (\nu + \mu) S_P = 0 \qquad (19)$$

$$\frac{dV_1}{dt} = \nu S_P - (\omega + \rho_1 + \mu) V_1 = 0 \qquad (20)$$

$$\frac{dV_2}{dt} = \omega \mathrm{V}_1 - (\omega_1 + \rho_2 + \mu) V_2 = 0 \qquad (21)$$

$$\frac{dV_3}{dt} = \omega_1 \mathrm{V}_2 - (\rho_3 + \mu) V_3 = 0 \qquad (22)$$

$$\frac{dS_{S_1}}{dt} = \rho_1 \mathrm{V}_1 - \mu S_{S_1} = 0 \qquad (23)$$

$$\frac{dS_{S_2}}{dt} = \rho_2 V_2 - \mu S_{S_2} = 0 \qquad (24)$$

$$\frac{dS_{S_3}}{dt} = \rho_3 V_3 - \mu S_{S_3} = 0 \qquad (25)$$

The solution for the disease-free equilibrium is given from (26) to (32):

$$S_P^* = \frac{\mu}{\nu + \mu} \qquad (26)$$

$$V_1^* = \frac{\nu}{\omega + \rho_1 + \mu} \cdot \frac{\mu}{\mu + \nu} \qquad (27)$$

$$V_2^* = \frac{\omega}{\omega_1 + \rho_2 + \mu} \cdot \frac{\nu}{\omega + \rho_1 + \mu} \cdot \frac{\mu}{\mu + \nu} \qquad (28)$$

$$V_3^* = \frac{\omega_1}{(\rho_3 + \mu)} \cdot \frac{\omega}{(\omega_1 + \rho_2 + \mu)} \cdot \frac{\nu}{\omega + \rho_1 + \mu} \cdot \frac{\mu}{\mu + \nu} \qquad (29)$$

$$S_{S_1}^* = \frac{\rho_1}{\mu} V_1^* = \frac{\rho_1}{\omega + \rho_1 + \mu} \cdot \frac{\nu}{\mu + \nu} \qquad (30)$$

$$S_{S_2}^* = \frac{\rho_2}{\omega_1 + \rho_2 + \mu} \cdot \frac{\omega}{\omega + \rho_1 + \mu} \cdot \frac{\nu}{\mu + \nu} \qquad (31)$$

$$S_{S_3}^* = \frac{\rho_3}{(\rho_3 + \mu)} \cdot \frac{\omega_1}{(\omega_1 + \rho_2 + \mu)} \cdot \frac{\omega}{\omega + \rho_1 + \mu} \cdot \frac{\nu}{\mu + \nu} \qquad (32)$$

The reproduction number for the transmission dynamics system is determined by the formula (33):

$$
\begin{aligned}
\Re &= \frac{\beta}{\gamma+\mu}\left[S_P + \varepsilon S_S + \varepsilon_{V_1}V_1 + \varepsilon_{V_2}V_2 + \varepsilon_{V_3}V_3 + \varepsilon_1 S_{S_1} + \varepsilon_2 S_{S_2} + \varepsilon_3 S_{S_3}\right] \\
&= \frac{\beta}{\gamma+\mu}\left\{
\begin{array}{l}
\dfrac{\mu}{\nu+\mu}\left[\begin{array}{l} 1+\dfrac{\nu\varepsilon_{V_1}}{\omega+\rho_1+\mu}+\dfrac{\nu\varepsilon_{V_2}}{\omega+\rho_1+\mu}\dfrac{\omega}{\omega_1+\rho_2+\mu}+ \\ \dfrac{\nu\varepsilon_{V_3}}{\omega+\rho_1+\mu}\dfrac{\omega}{\omega_1+\rho_2+\mu}\dfrac{\omega_1}{\rho_3+\mu} \end{array}\right]+ \\
\dfrac{\nu}{\nu+\mu}\left[\begin{array}{l} \dfrac{\rho_1\varepsilon_1}{\omega+\rho_1+\mu}+\dfrac{\rho_2\varepsilon_2}{\omega+\rho_1+\mu}\dfrac{\omega}{\omega_1+\rho_2+\mu}+ \\ \dfrac{\rho_3\varepsilon_3}{\omega+\rho_1+\mu}\dfrac{\omega}{\omega_1+\rho_2+\mu}\dfrac{\omega_1}{\rho_3+\mu} \end{array}\right]
\end{array}
\right\}
\end{aligned}
\tag{33}
$$

The partial first-order differential concerning the administration rate of dose 1 is given by (34):

$$
\frac{\partial\Re}{\partial\nu} = \frac{\beta}{\gamma+\mu}\left\{
\begin{array}{l}
\dfrac{-\mu}{(\nu+\mu)^2}\left[\begin{array}{l} 1+\dfrac{\nu\varepsilon_{V_1}}{\omega+\rho_1+\mu}+\dfrac{\nu\varepsilon_{V_2}}{\omega+\rho_1+\mu}\dfrac{\omega}{\omega_1+\rho_2+\mu}+ \\ \dfrac{\nu\varepsilon_{V_3}}{\omega+\rho_1+\mu}\dfrac{\omega}{\omega_1+\rho_2+\mu}\dfrac{\omega_1}{\rho_3+\mu} \end{array}\right]+ \\
\dfrac{\mu}{(\nu+\mu)^2}\left[\begin{array}{l} \dfrac{\rho_1\varepsilon_1}{\omega+\rho_1+\mu}+\dfrac{\rho_2\varepsilon_2}{\omega+\rho_1+\mu}\dfrac{\omega}{\omega_1+\rho_2+\mu}+ \\ \dfrac{\rho_3\varepsilon_3}{\omega+\rho_1+\mu}\dfrac{\omega}{\omega_1+\rho_2+\mu}\dfrac{\omega_1}{\rho_3+\mu} \end{array}\right]
\end{array}
\right\}
\tag{34}
$$

The sign for $\frac{\partial R}{\partial \nu}$ is indeterministic and hinges on multiple factors including the rollout rate of doses, rate of immunity waning, infection rate after vaccination, rate of birth/death, and so on. For the reproduction number to be a monotonic decreasing function of the vaccination rate of dose 1 (i.e., $\frac{\partial R}{\partial \nu} < 0$), the condition in (35) needs to be satisfied:

$$(\omega_1+\rho_2+\mu)(\rho_3+\mu)(\rho_1\varepsilon_1-\nu\varepsilon_{V_1})+(\rho_3+\mu)\omega(\rho_2\varepsilon_2-\nu\varepsilon_{V_2})+\omega\omega_1(\rho_3\varepsilon_3-\nu\varepsilon_{V_3})< (\omega+\rho_1+\mu)(\omega_1+\rho_2+\mu)(\rho_3+\mu) \qquad (35)$$

After simplification, the condition as stated in formula (36) need to be satisfied:

$$\frac{(\rho_1\varepsilon_1-\nu\varepsilon_{V_1})}{(\omega+\rho_1+\mu)}+\frac{\omega(\rho_2\varepsilon_2-\nu\varepsilon_{V_2})}{(\omega+\rho_1+\mu)(\omega_1+\rho_2+\mu)}+\frac{\omega\omega_1(\rho_3\varepsilon_3-\nu\varepsilon_{V_3})}{(\omega+\rho_1+\mu)(\omega_1+\rho_2+\mu)(\rho_3+\mu)}<1 \qquad (36)$$

Following the similar vein as dose 1, derive the relative relation over the administration rate of dose 2:

$$\frac{\partial\Re}{\partial\omega}=\frac{\beta}{\gamma+\mu}\left\{\begin{matrix}\frac{\mu}{\nu+\mu}\left[\begin{matrix}\frac{-\nu\varepsilon_{V_1}}{(\omega+\rho_1+\mu)^2}+\frac{\nu\varepsilon_{V_2}}{(\omega+\rho_1+\mu)^2}\frac{\rho_1+\mu}{\omega_1+\rho_2+\mu}+\\ \frac{\nu\varepsilon_{V_3}}{(\omega+\rho_1+\mu)^2}\frac{\rho_1+\mu}{\omega_1+\rho_2+\mu}\frac{\omega_1}{\rho_3+\mu}\end{matrix}\right]+\\ \frac{\nu}{\nu+\mu}\left[\begin{matrix}\frac{-\rho_1\varepsilon_1}{(\omega+\rho_1+\mu)^2}+\frac{\rho_2\varepsilon_2}{(\omega+\rho_1+\mu)^2}\frac{\rho_1+\mu}{\omega_1+\rho_2+\mu}+\\ \frac{\rho_3\varepsilon_3}{(\omega+\rho_1+\mu)^2}\frac{\rho_1+\mu}{\omega_1+\rho_2+\mu}\frac{\omega_1}{\rho_3+\mu}\end{matrix}\right]\end{matrix}\right\} \qquad (37)$$

For R to be a monotonic decreasing function of dose 2 administration rate, it needs to satisfy the condition in formula (38):

$$(\mu\varepsilon_{V_2}+\rho_2\varepsilon_2)(\rho_1+\mu)(\rho_3+\mu)+(\mu\varepsilon_{V_3}+\rho_3\varepsilon_3)(\rho_1+\mu)\omega_1< (\mu\varepsilon_{V_1}+\rho_1\varepsilon_1)(\omega_1+\rho_2+\mu)(\rho_3+\mu) \qquad (38)$$

And after simplification, obtain the condition of (39):

$$\frac{\mu\varepsilon_{V_2}+\rho_2\varepsilon_2}{(\rho_2+\mu)(\omega_1+\rho_2+\mu)}+\frac{(\mu\varepsilon_{V_3}+\rho_3\varepsilon_3)\omega_1}{(\rho_3+\mu)(\rho_2+\mu)(\omega_1+\rho_2+\mu)}<\frac{\mu\varepsilon_{V_1}+\rho_1\varepsilon_1}{(\rho_1+\mu)(\rho_2+\mu)} \tag{39}$$

Generalize the analysis to dose 3:

$$\frac{\partial\Re}{\partial\omega_1}=\frac{\beta}{\gamma+\mu}\left\{\begin{array}{l}\dfrac{\mu}{\nu+\mu}\left[\begin{array}{l}\dfrac{\nu\varepsilon_{V_2}}{\omega+\rho_1+\mu}\dfrac{-\omega}{(\omega_1+\rho_2+\mu)^2}+\\ \dfrac{\nu\varepsilon_{V_3}}{\omega+\rho_1+\mu}\dfrac{\omega}{(\omega_1+\rho_2+\mu)^2}\dfrac{\rho_2+\mu}{\rho_3+\mu}\end{array}\right]+\\ \dfrac{\nu}{\nu+\mu}\left[\begin{array}{l}\dfrac{\rho_2\varepsilon_2}{\omega+\rho_1+\mu}\dfrac{-\omega}{(\omega_1+\rho_2+\mu)^2}+\\ \dfrac{\rho_3\varepsilon_3}{\omega+\rho_1+\mu}\dfrac{\omega}{(\omega_1+\rho_2+\mu)^2}\dfrac{\rho_2+\mu}{\rho_3+\mu}\end{array}\right]\end{array}\right\} \tag{40}$$

For R to be a decreasing function of administration rate, i.e., $\frac{\partial\Re}{\partial\omega_1}<0$, obtain the condition depicted in (41):

$$\frac{\mu\varepsilon_{V_3}+\rho_3\varepsilon_3}{\rho_3+\mu}<\frac{\mu\varepsilon_{V_2}+\rho_2\varepsilon_2}{\rho_2+\mu} \tag{41}$$

### Supplemental Figures

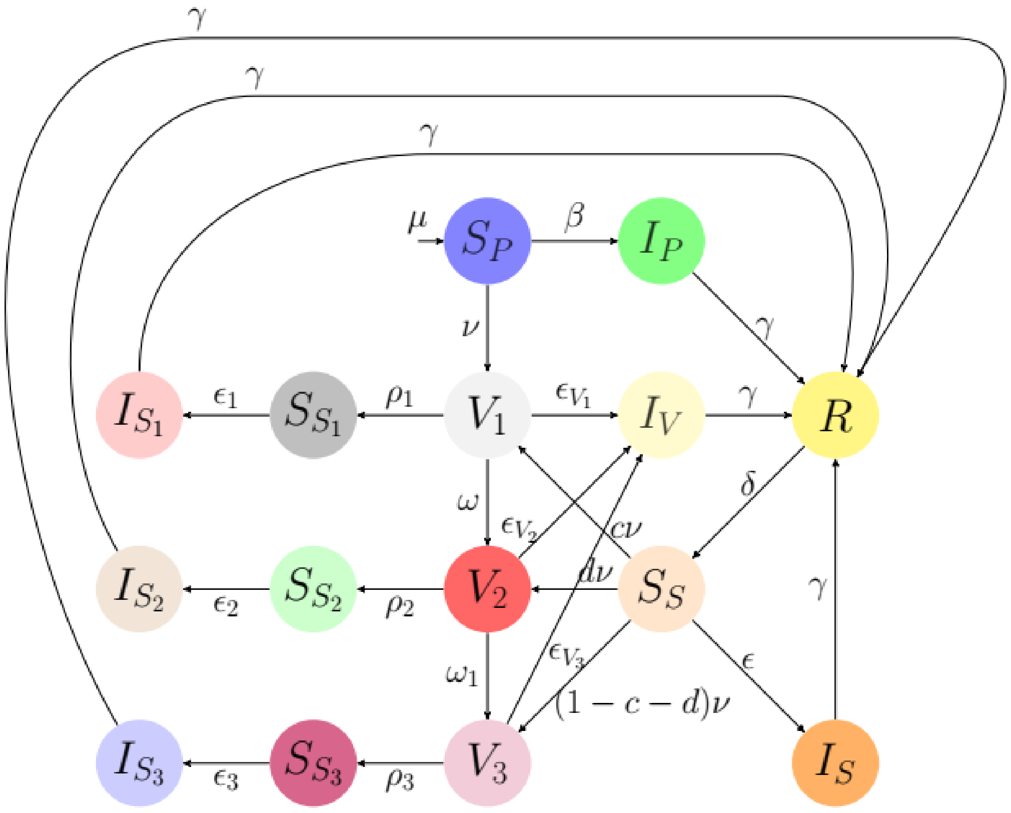


Figure S1: The framework of COVID-19 transmission dynamics

- Dose 1 vaccination
- Dose-1 immunity wanes
- Infection after dose-1 immunity wanes
- Dose 2 vaccination
- Dose-2 immunity wanes
- Primary infection
- Infection after vaccination
- Recovered
- Partial susceptibility
- Secondary infection
- Dose 3 vaccination
- Dose-3 immunity wanes
- Infection after dose-3 immunity wanes
- Full susceptibility
- Infection after dose-2 immunity wanes

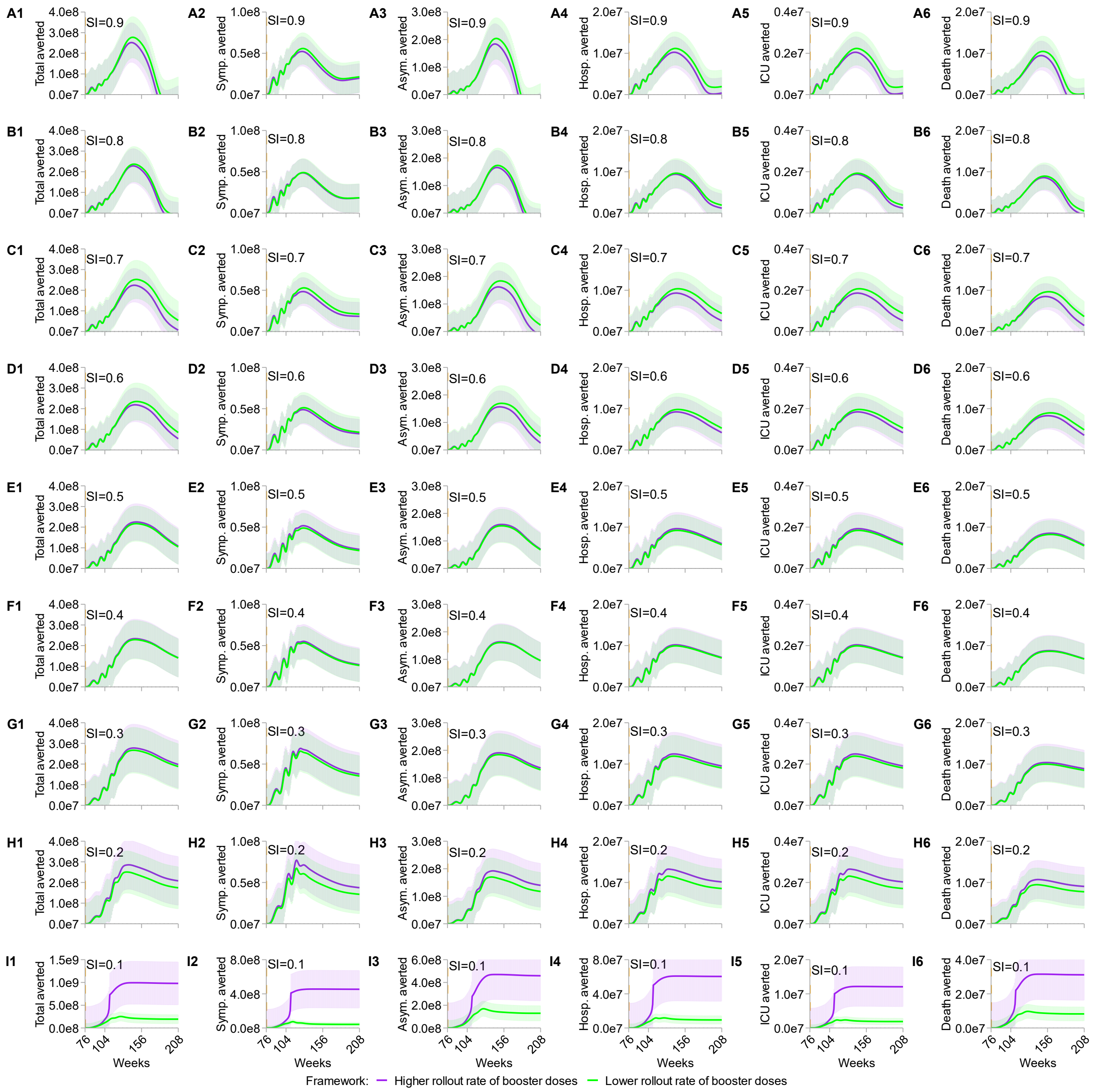


**Figure S2: Sensitivity test on the rate of waning to susceptibility after primary dose 1 vaccination for stratified cases averted of six disease burdens.** SI varies from 0.9 (A1 to A6) to 0.1 (I1 to I6) in 0.1 discrete increments. All scenarios denote the cumulative cases averted over four years and the population size is equal to one billion. "Total averted" represents the total infections averted; "Symp. averted" depicts the symptomatic cases averted; "Asym. averted" represents the asymptomatic cases averted; "Hosp. averted" depicts the hospitalization cases averted; "ICU averted" outlines the averted cases of patients needing intensive care unit; "Death averted" delineates the death cases averted. For each SI scenario, higher (purple curves and shaded areas) and lower (green curves and shaded areas) rollout rates of booster doses are equal to 4 and 0.5 per year respectively. Shaded light colors sketch the 95% confidence intervals of the simulations.

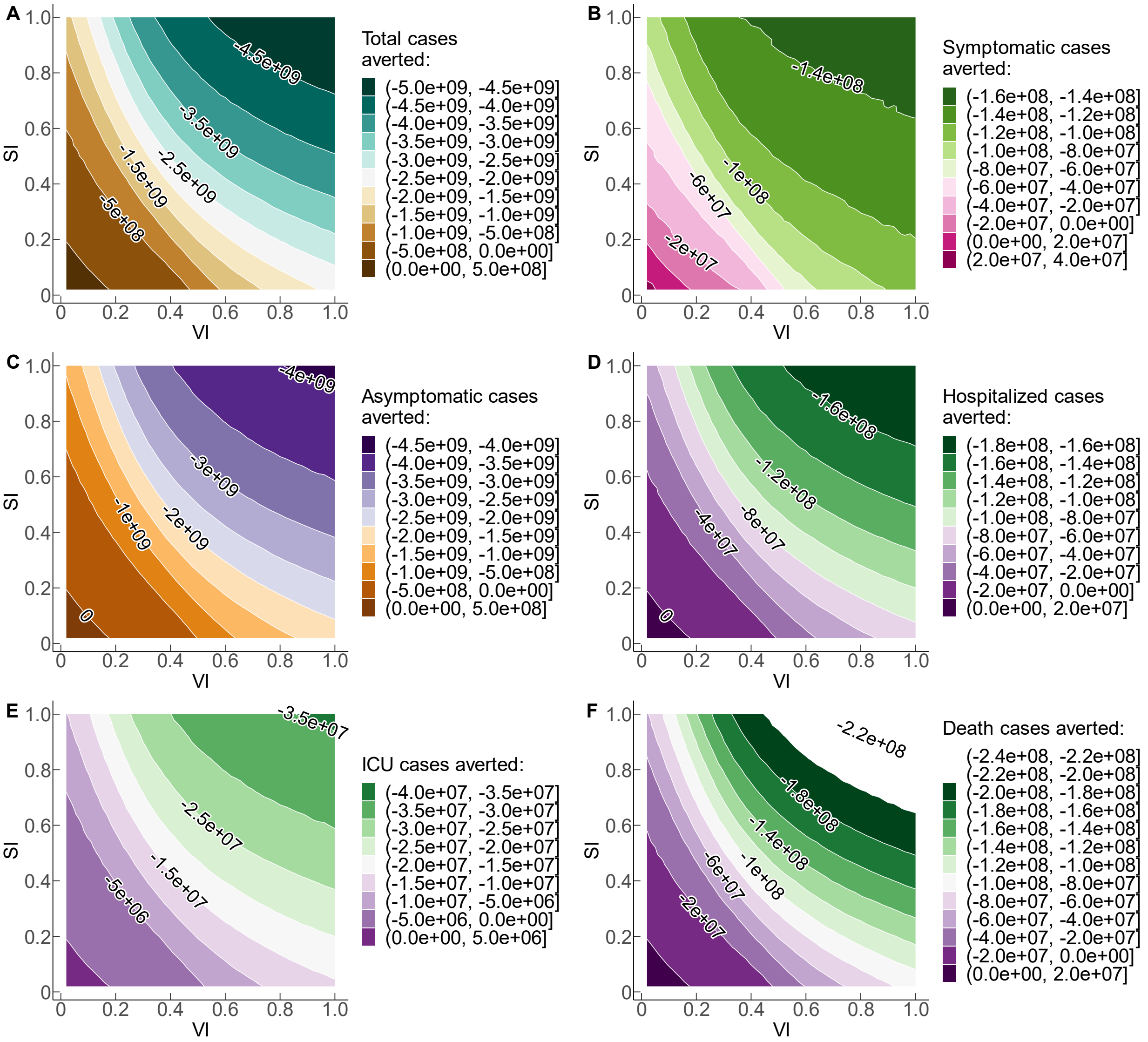


**Figure S3: Trade-offs sensitivity test for stratified disease burdens with the rate of waning to susceptibility after primary dose 1 vaccination.** Sensitivity tests of the cumulative stratified cases averted using the rate of waning to susceptibility after primary dose 1 vaccination with SI and VI ranging from 0 to 1.0 using 50 simulations for each metric respectively. All scenarios denote the cumulative cases averted over four years and the population size is equal to one billion. Positive numbers on each curve illustrate the positive aversion effect, and negative numbers depict the negative trade-off (i.e., negative cases averted) for each health outcome when VI and SI are sub-optimal ceteris paribus.

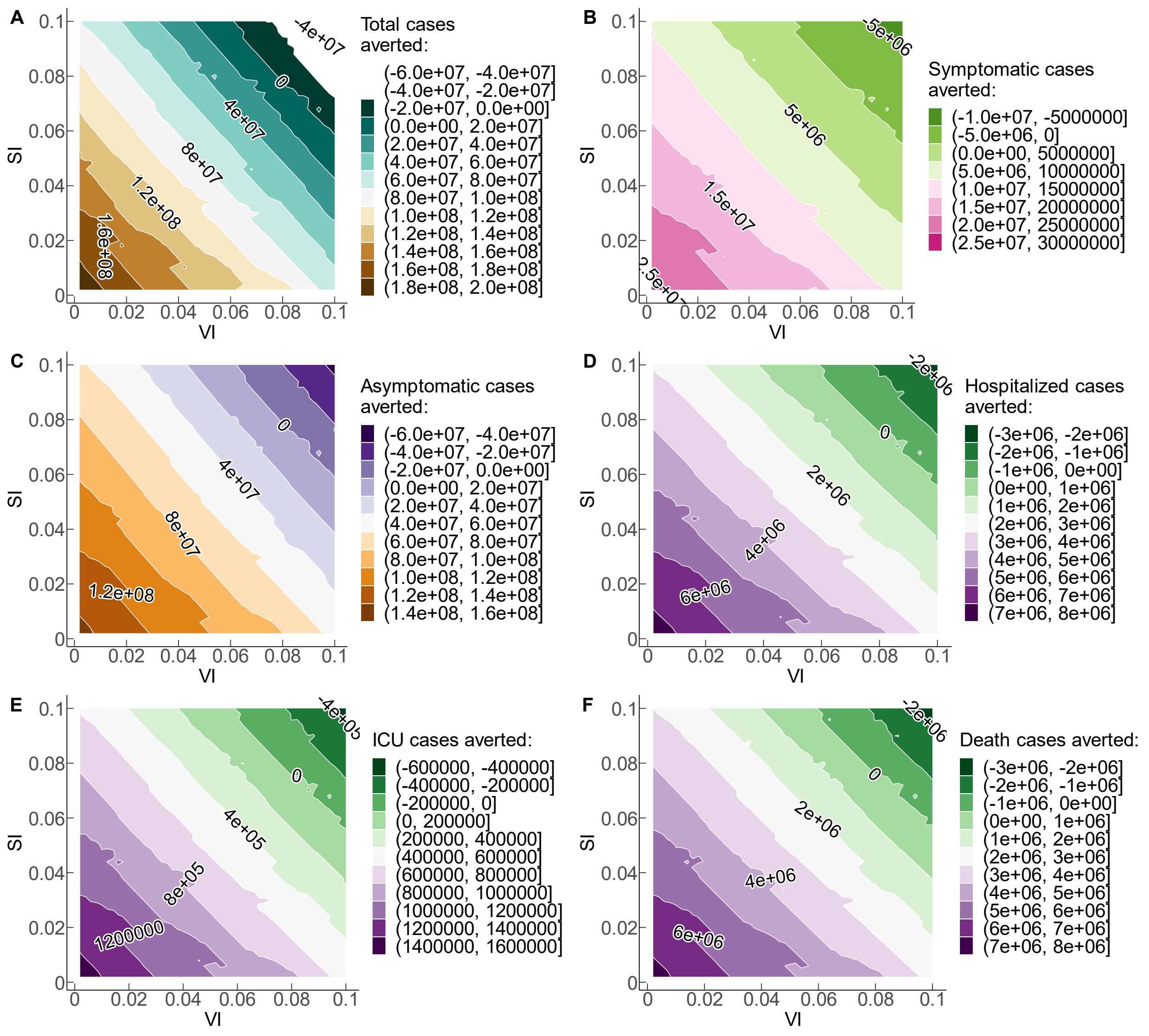


**Figure S4: Trade-offs sensitivity test for stratified disease burdens with the rate of waning to susceptibility after primary dose 1 vaccination.** Trade-off sensitivity tests of the cumulative stratified cases averted using the rate of waning to susceptibility after primary dose 1 vaccination with SI and VI ranging from 0 to 0.1 using 50 simulations for each metric respectively. All scenarios denote the cumulative cases averted over four years and the population size is equal to one billion. Positive numbers on each curve illustrate the positive aversion effect, and negative numbers depict the negative trade-off (i.e., negative cases averted) for each health outcome when VI and SI are sub-optimal ceteris paribus.

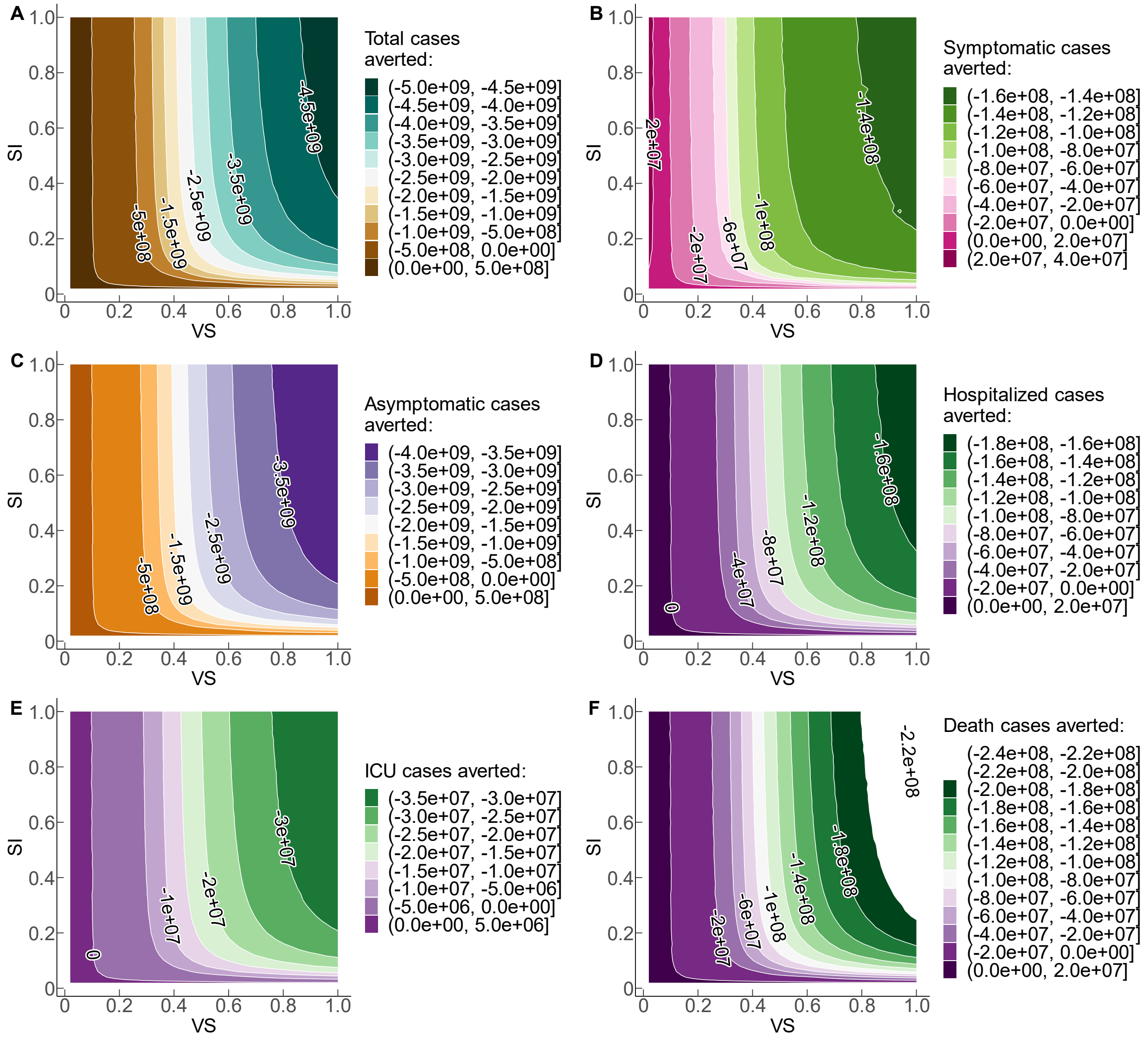


**Figure S5: Trade-offs sensitivity test for stratified disease burdens with the rate of waning to susceptibility after primary dose 1 vaccination.** Sensitivity tests of the cumulative stratified cases averted using the rate of waning to susceptibility after primary dose 1 vaccination with SI and VS ranging from 0 to 1.0 using 50 simulations for each metric respectively. All scenarios denote the cumulative cases averted over four years and the population size is equal to one billion. Positive numbers on each curve illustrate the positive aversion effect, and negative numbers depict the negative trade-off (i.e., negative cases averted) for each health outcome when VI and SI are sub-optimal ceteris paribus.

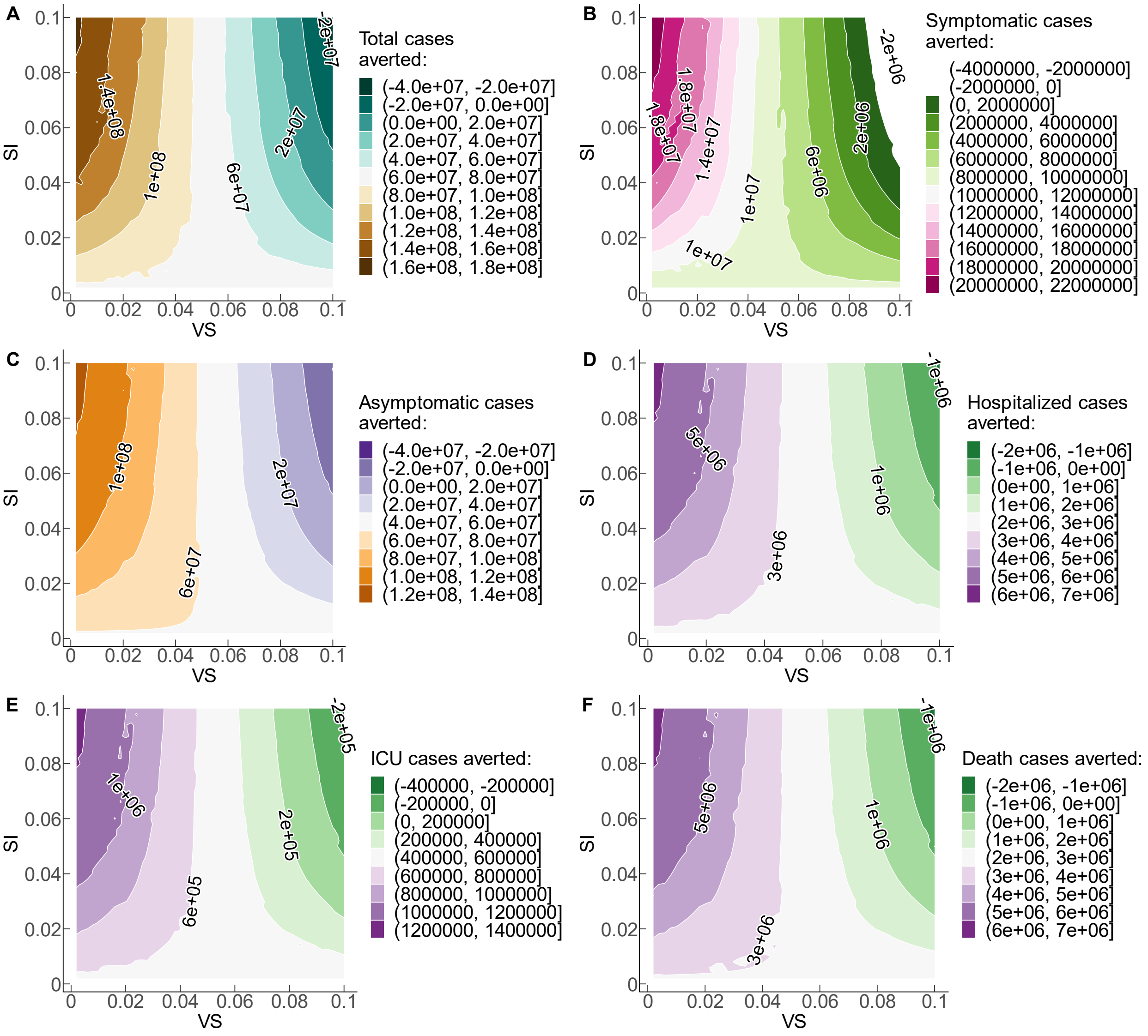


**Figure S6: Trade-offs sensitivity test for stratified disease burdens with the rate of waning to susceptibility after primary dose 1 vaccination.** Sensitivity tests of the cumulative stratified cases averted using the rate of waning to susceptibility after primary dose 1 vaccination with SI and VS ranging from 0 to 0.1 using 50 simulations for each metric respectively. All scenarios denote the cumulative cases averted over four years and the population size is equal to one billion. Positive numbers on each curve illustrate the positive aversion effect, and negative numbers depict the negative trade-off (i.e., negative cases averted) for each health outcome when VI and SI are sub-optimal ceteris paribus.

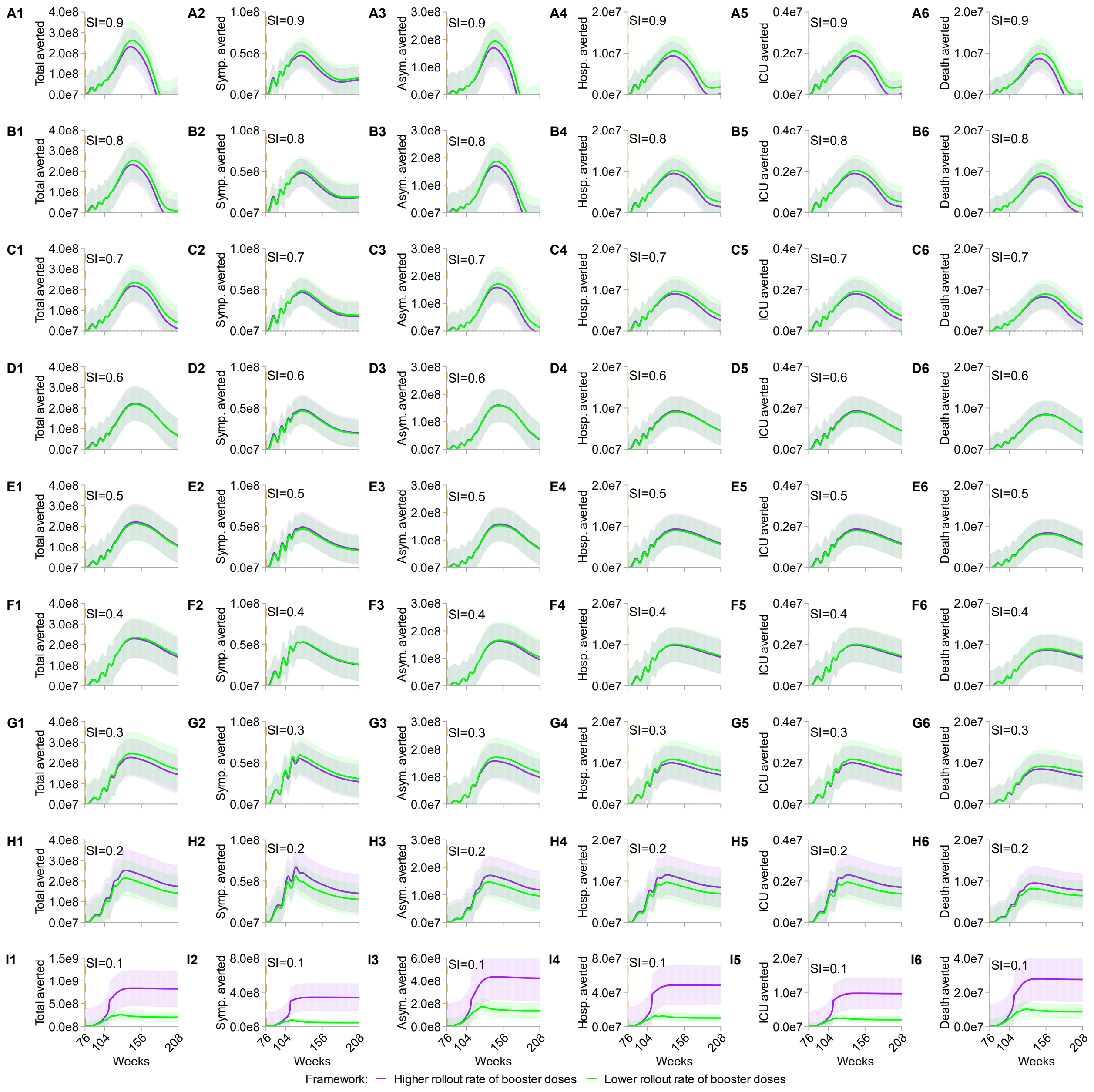


**Figure S7: Sensitivity test on the rate of waning to susceptibility after primary dose 2 vaccination for stratified cases averted of disease burdens.** SI varies from 0.9 (A1 to A6) to 0.1 (I1 to I6) in 0.1 increments. All scenarios denote the cumulative cases averted over four years and the population size is equal to one billion. "Total averted" represents the total infections averted; "Symp. averted" depicts the symptomatic cases averted; "Asym. averted" represents the asymptomatic cases averted; "Hosp. averted" depicts the hospitalization cases averted; "ICU averted" outlines the averted cases of patients needing intensive care unit; "Death averted" delineates the death cases averted. For each SI scenario, higher (purple curves and shaded areas) and lower (green curves and shaded areas) rollout rates of booster doses are equal to 4 and 0.5 per year respectively. Shaded light colors sketch the 95% confidence intervals of the simulations.

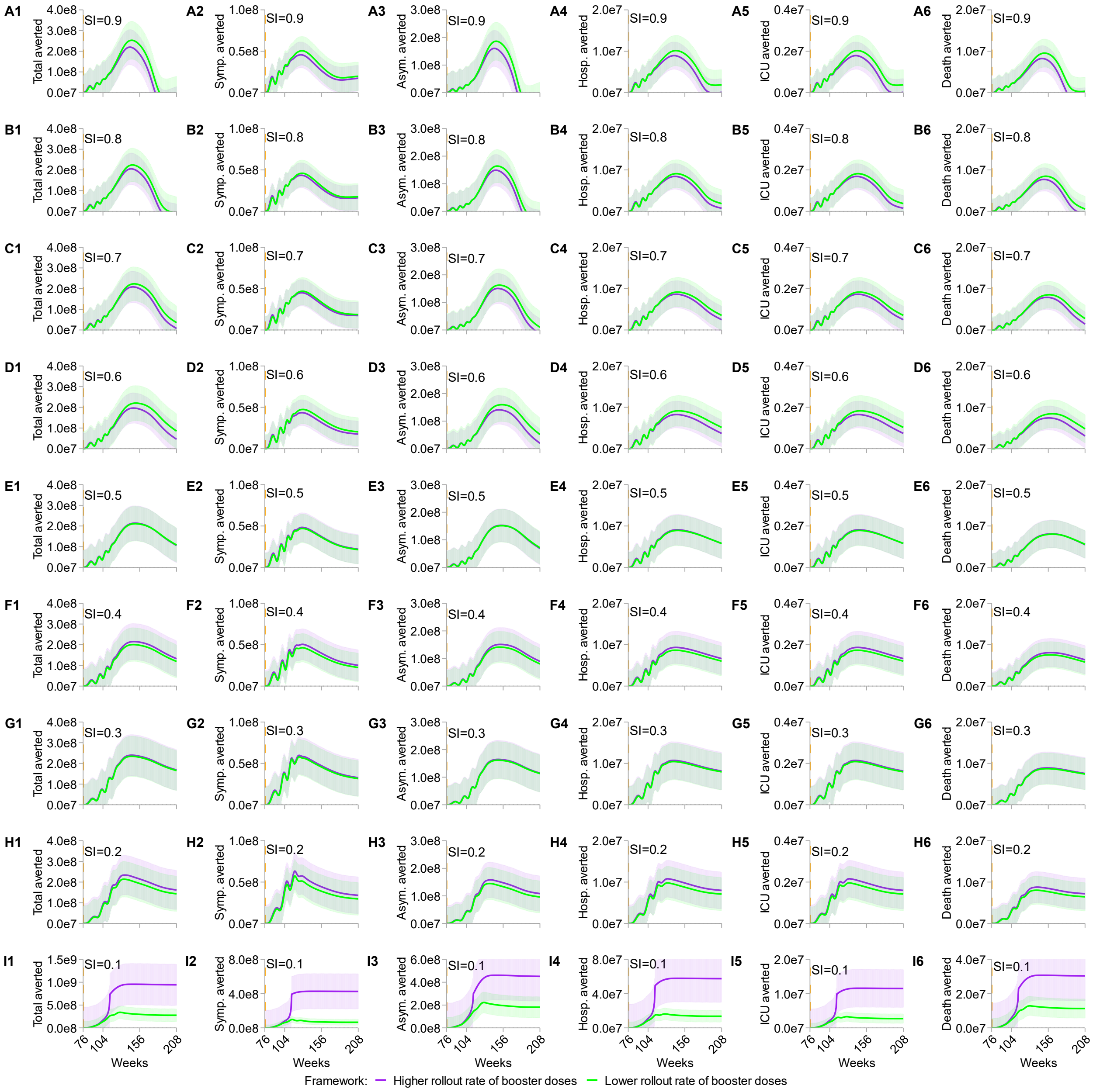


**Figure S8: Sensitivity test on the rate of birth/death (lower rate) for stratified metrics of disease burdens.** SI varies from 0.9 (A1 to A6) to 0.1 (I1 to I6) in 0.1 increments. All scenarios denote the cumulative cases averted over four years and the population size is equal to one billion. “Total averted” represents the total infections averted; “Symp. averted” depicts the symptomatic cases averted; “Asym. averted” represents the asymptomatic cases averted; “Hosp. averted” depicts the hospitalization cases averted; “ICU averted” outlines the averted cases of patients needing intensive care unit; “Death averted” delineates the death cases averted. For each SI scenario, higher (purple curves and shaded areas) and lower (green curves and shaded areas) rollout rates of booster doses are equal to 4 and 0.5 per year respectively. Shaded light colors sketch the 95% confidence intervals of the simulations.

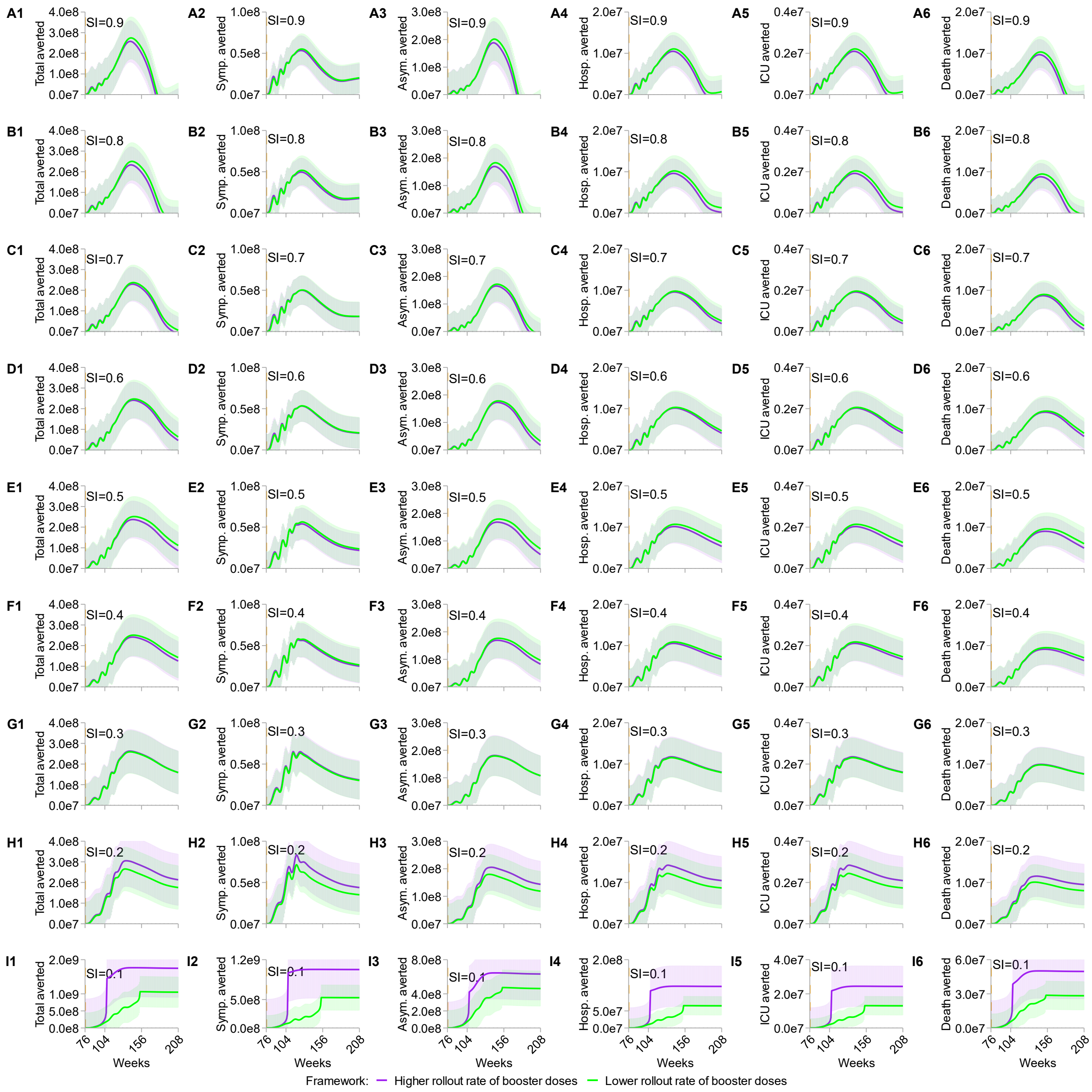


**Figure S9: Sensitivity test on the rate of birth/death (greater rate) for stratified metrics of disease burdens.** SI varies from 0.9 (A1 to A6) to 0.1 (I1 to I6) in 0.1 increments. All scenarios denote the cumulative cases averted over four years and the population size is equal to one billion. "Total averted" represents the total cases averted; "Symp. averted" depicts the symptomatic cases averted; "Asym. averted" represents the asymptomatic cases averted; "Hosp. averted" depicts the hospitalization cases averted; "ICU averted" outlines the averted cases of patients needing intensive care unit; "Death averted" delineates the death cases averted. For each SI scenario, higher (purple curves and shaded areas) and lower (green curves and shaded areas) rollout rates of booster doses are equal to 4 and 0.5 per year respectively. Shaded light colors sketch the 95% confidence intervals of the simulations.

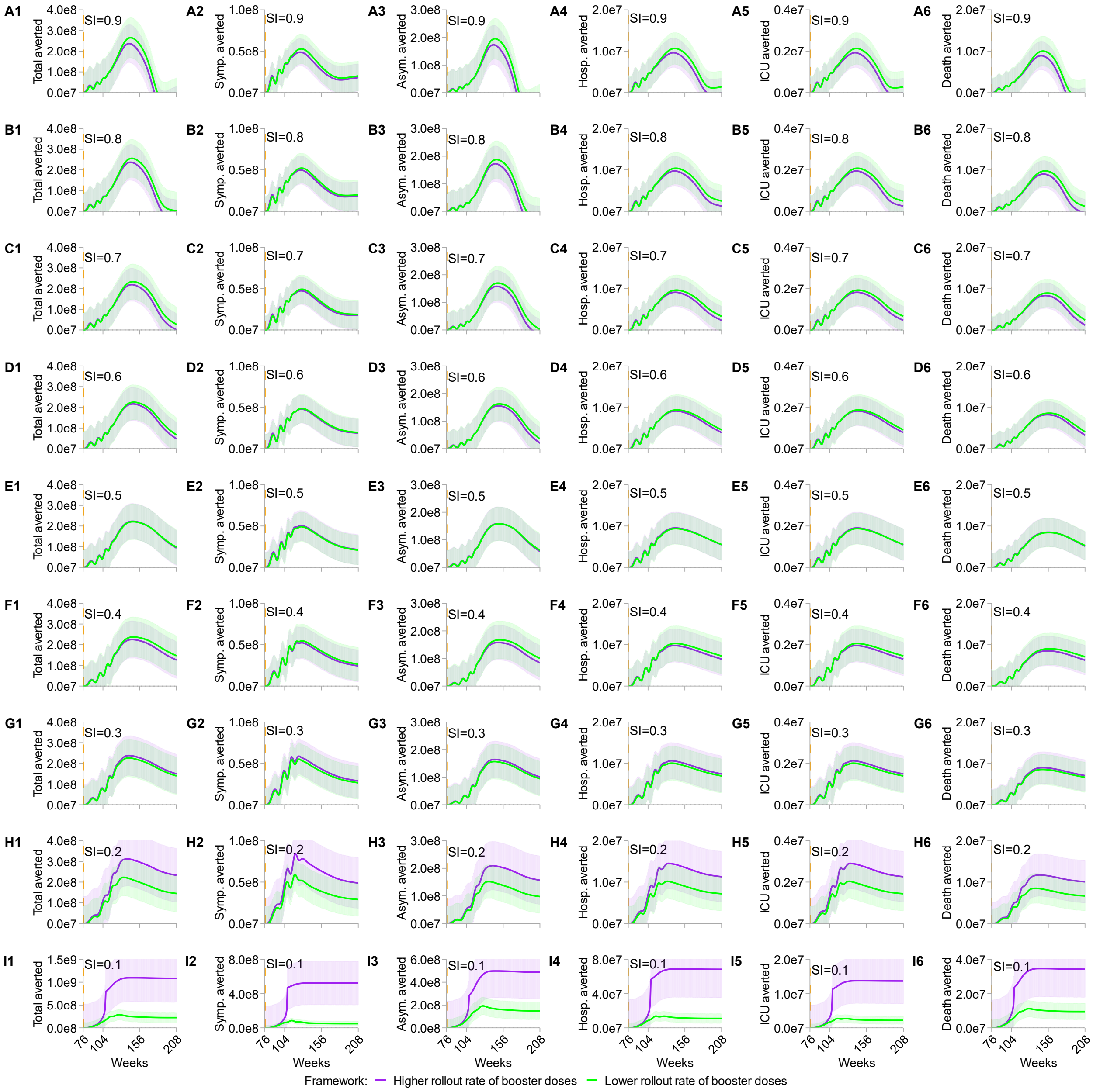


**Figure S10: Sensitivity test on rollout rate of primary dose 1 for stratified metrics of disease burdens.** SI varies from 0.9 (A1 to A6) to 0.1 (I1 to I6) in 0.1 increments. All scenarios denote the cumulative cases averted over four years and the population size is equal to one billion. "Total averted" represents the total infections averted; "Symp. averted" depicts the symptomatic cases averted; "Asym. averted" represents the asymptomatic cases averted; "Hosp. averted" depicts the hospitalization cases averted; "ICU averted" outlines the averted cases of patients needing intensive care unit; "Death averted" delineates the death cases averted. For each SI scenario, higher (purple curves and shaded areas) and lower (green curves and shaded areas) rollout rates of booster doses are equal to 4 and 0.5 per year respectively. Shaded light colors sketch the 95% confidence intervals of the simulations.

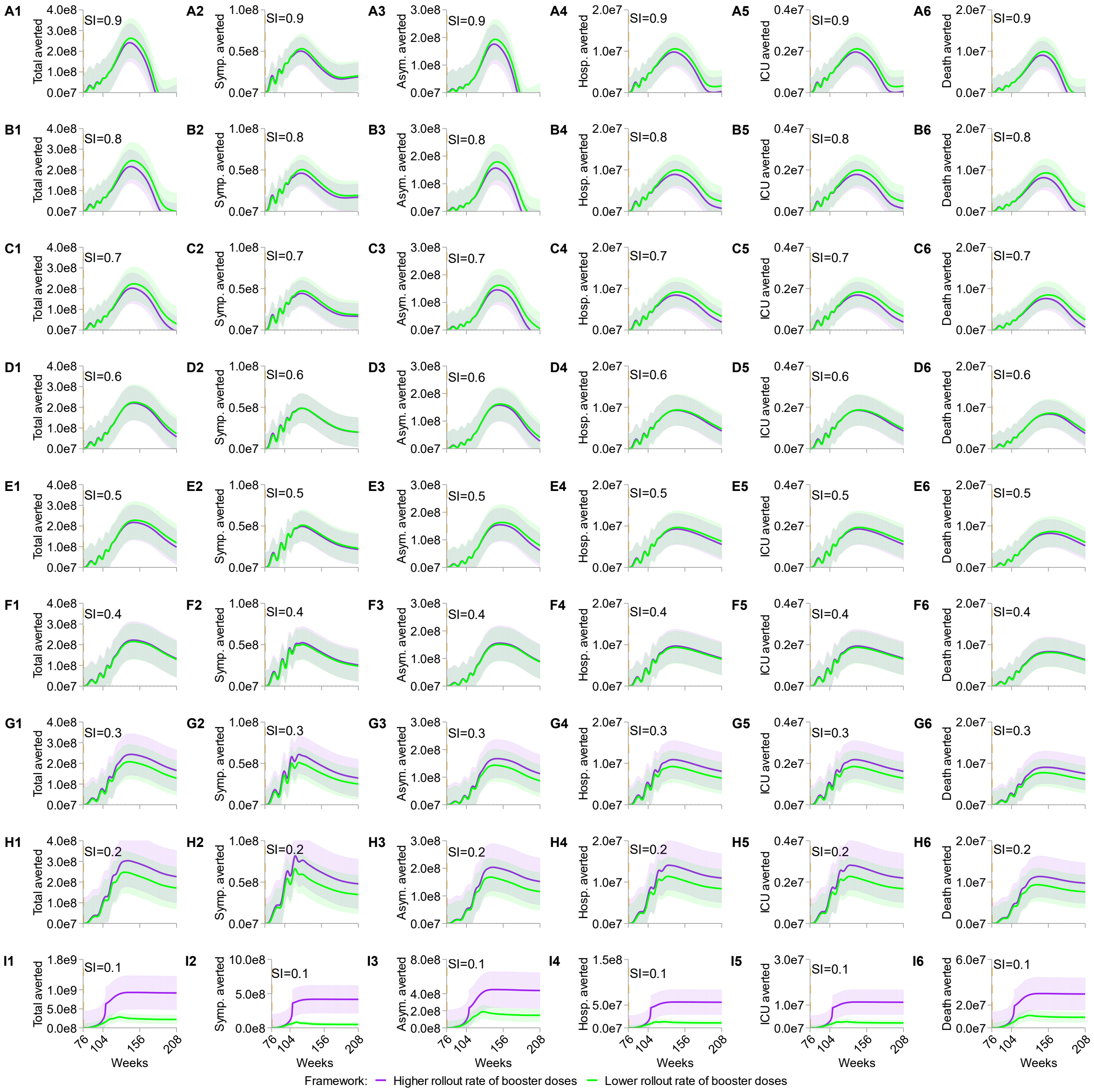


**Figure S11: Sensitivity test on rollout rate of primary dose 2 for stratified metrics of disease burdens.** SI varies from 0.9 (A1 to A6) to 0.1 (I1 to I6) in 0.1 increments. All scenarios denote the cumulative cases averted over four years and the population size is equal to one billion. “Total averted” represents the total cases averted; “Symp. averted” depicts the symptomatic cases averted; “Asym. averted” represents the asymptomatic cases averted; “Hosp. averted” depicts the hospitalization cases averted; “ICU averted” outlines the averted cases of patients needing intensive care unit; “Death averted” delineates the death cases averted. For each SI scenario, higher (purple curves and shaded areas) and lower (green curves and shaded areas) rollout rates of booster doses are equal to 4 and 0.5 per year respectively. Shaded light colors sketch the 95% confidence intervals of the simulations.

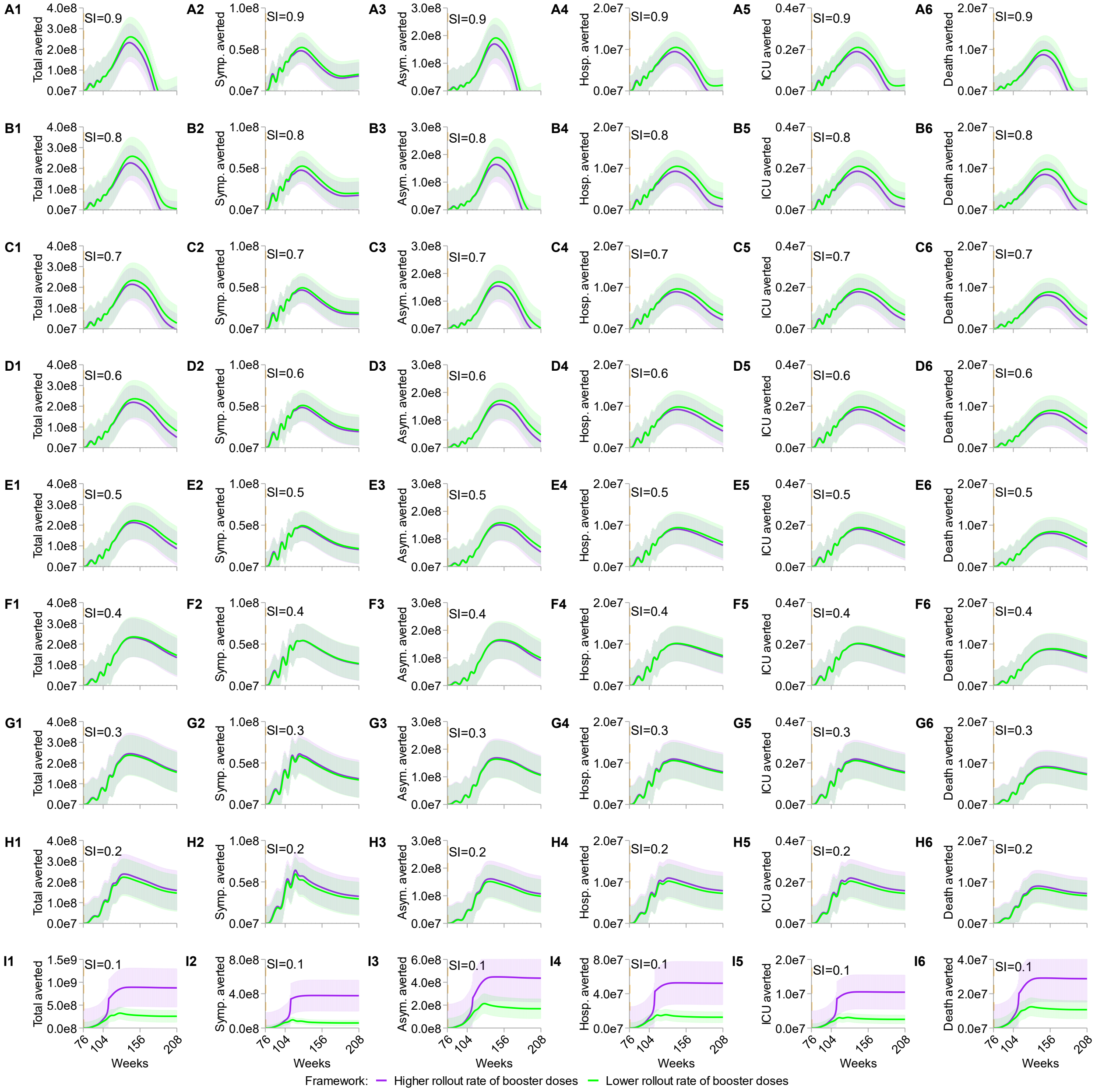


**Figure S12: Sensitivity test on rollout rate (greater rate) of booster doses for stratified metrics of disease burdens.** SI varies from 0.9 (A1 to A6) to 0.1 (I1 to I6) in 0.1 increments. All scenarios denote the cumulative cases averted over four years and the population size is equal to one billion. "Total averted" represents the total cases averted; "Symp. averted" depicts the symptomatic cases averted; "Asym. averted" represents the asymptomatic cases averted; "Hosp. averted" depicts the hospitalization cases averted; "ICU averted" outlines the averted cases of patients needing intensive care unit; "Death averted" delineates the death cases averted. For each SI scenario, higher (purple curves and shaded areas) and lower (green curves and shaded areas) rollout rates of booster doses are equal to 4 and 0.5 per year respectively. Shaded light colors sketch the 95% confidence intervals of the simulations.

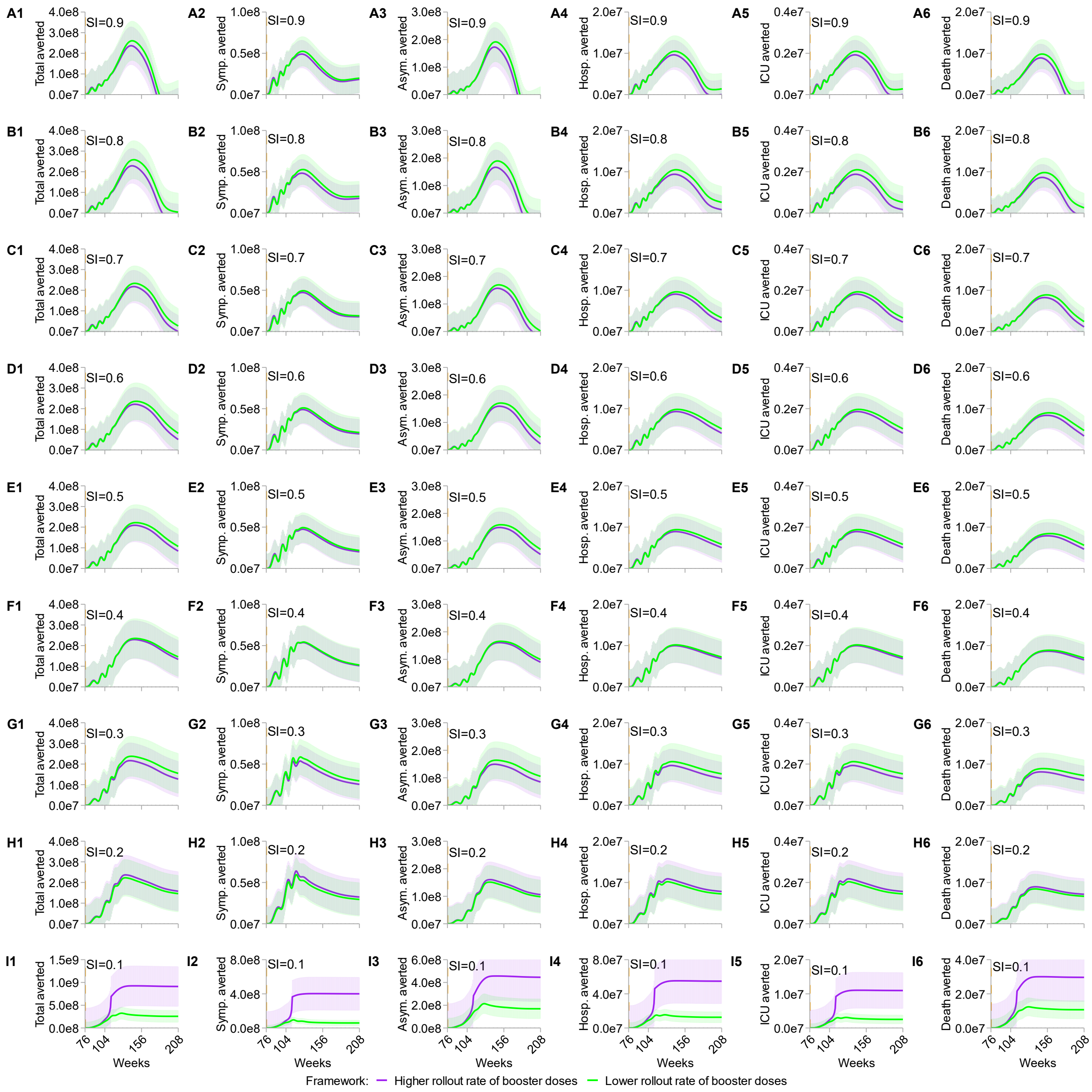

**Figure S13: Sensitivity test on rollout rate (lower rate) of booster doses for stratified metrics of disease burdens.** SI varies from 0.9 (A1 to A6) to 0.1 (I1 to I6) in 0.1 increments. All scenarios denote the cumulative cases averted over four years and the population size is equal to one billion. “Total averted” represents the total cases averted; “Symp. averted” depicts the symptomatic cases averted; “Asym. averted” represents the asymptomatic cases averted; “Hosp. averted” depicts the hospitalization cases averted; “ICU averted” outlines the averted cases of patients needing intensive care unit; “Death averted” delineates the death cases averted. For each SI scenario, higher (purple curves and shaded areas) and lower (green curves and shaded areas) rollout rates of booster doses are equal to 4 and 0.5 per year respectively. Shaded light colors sketch the 95% confidence intervals of the simulations.

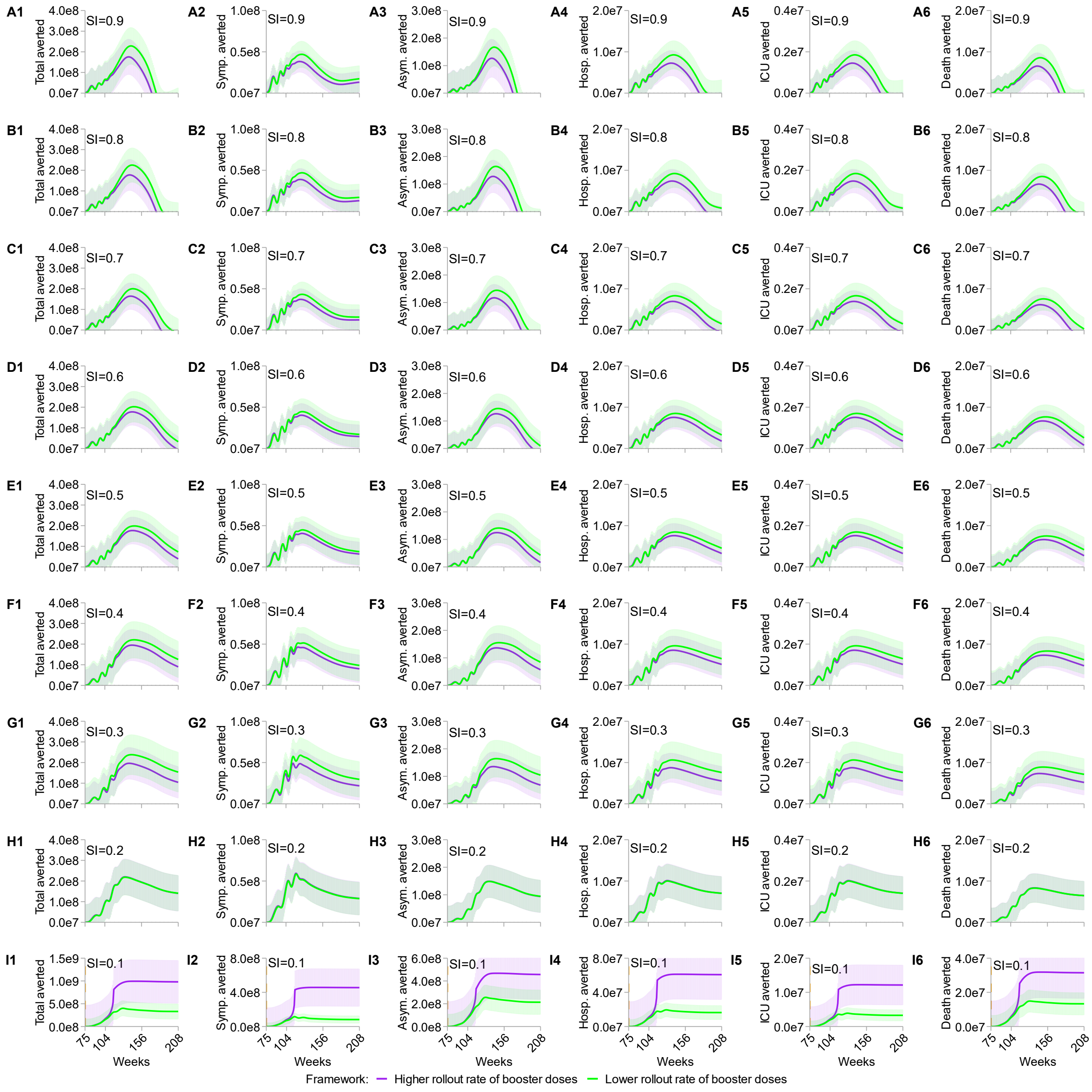


**Figure S14: Sensitivity test on vaccination timing of booster doses (earlier) for stratified metrics of disease burdens.** SI varies from 0.9 (A1 to A6) to 0.1 (I1 to I6) in 0.1 increments. All scenarios denote the cumulative cases averted over four years and the population size is equal to one billion. "Total averted" represents the total cases averted; "Symp. averted" depicts the symptomatic cases averted; "Asym. averted" represents the asymptomatic cases averted; "Hosp. averted" depicts the hospitalization cases averted; "ICU averted" outlines the averted cases of patients needing intensive care unit; "Death averted" delineates the death cases averted. For each SI scenario, higher (purple curves and shaded areas) and lower (green curves and shaded areas) rollout rates of booster doses are equal to 4 and 0.5 per year respectively. Shaded light colors sketch the 95% confidence intervals of the simulations.

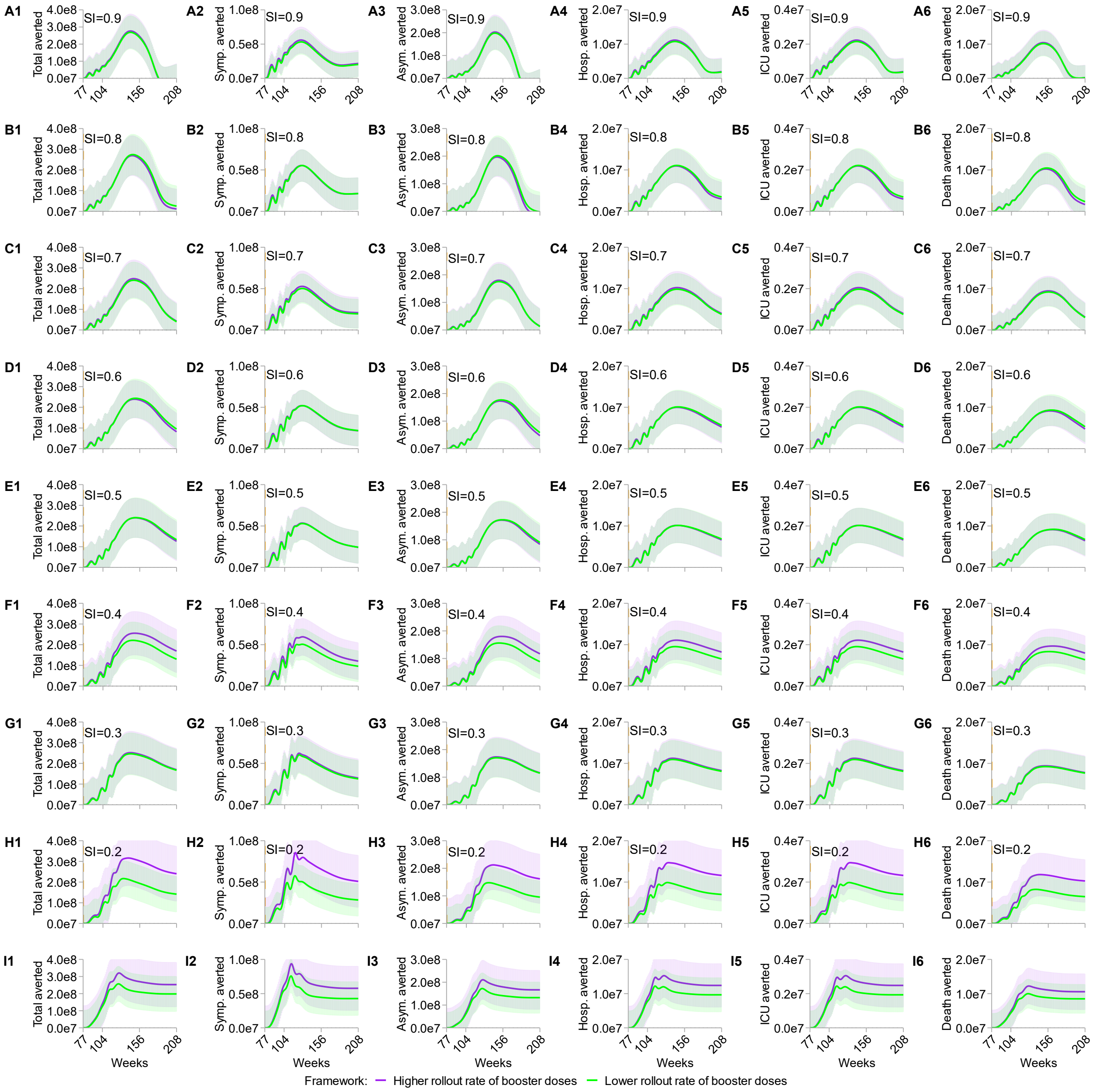


**Figure S15: Sensitivity test on vaccination timing of booster doses (delayed) for stratified metrics of disease burdens.** SI varies from 0.9 (A1 to A6) to 0.1 (I1 to I6) in 0.1 increments. All scenarios denote the cumulative cases averted over four years and the population size is equal to one billion. "Total averted" represents the total cases averted; "Symp. averted" depicts the symptomatic cases averted; "Asym. averted" represents the asymptomatic cases averted; "Hosp. averted" depicts the hospitalization cases averted; "ICU averted" outlines the averted cases of patients needing intensive care unit; "Death averted" delineates the death cases averted. For each SI scenario, higher (purple curves and shaded areas) and lower (green curves and shaded areas) rollout rates of booster doses are equal to 4 and 0.5 per year respectively. Shaded light colors sketch the 95% confidence intervals of the simulations.

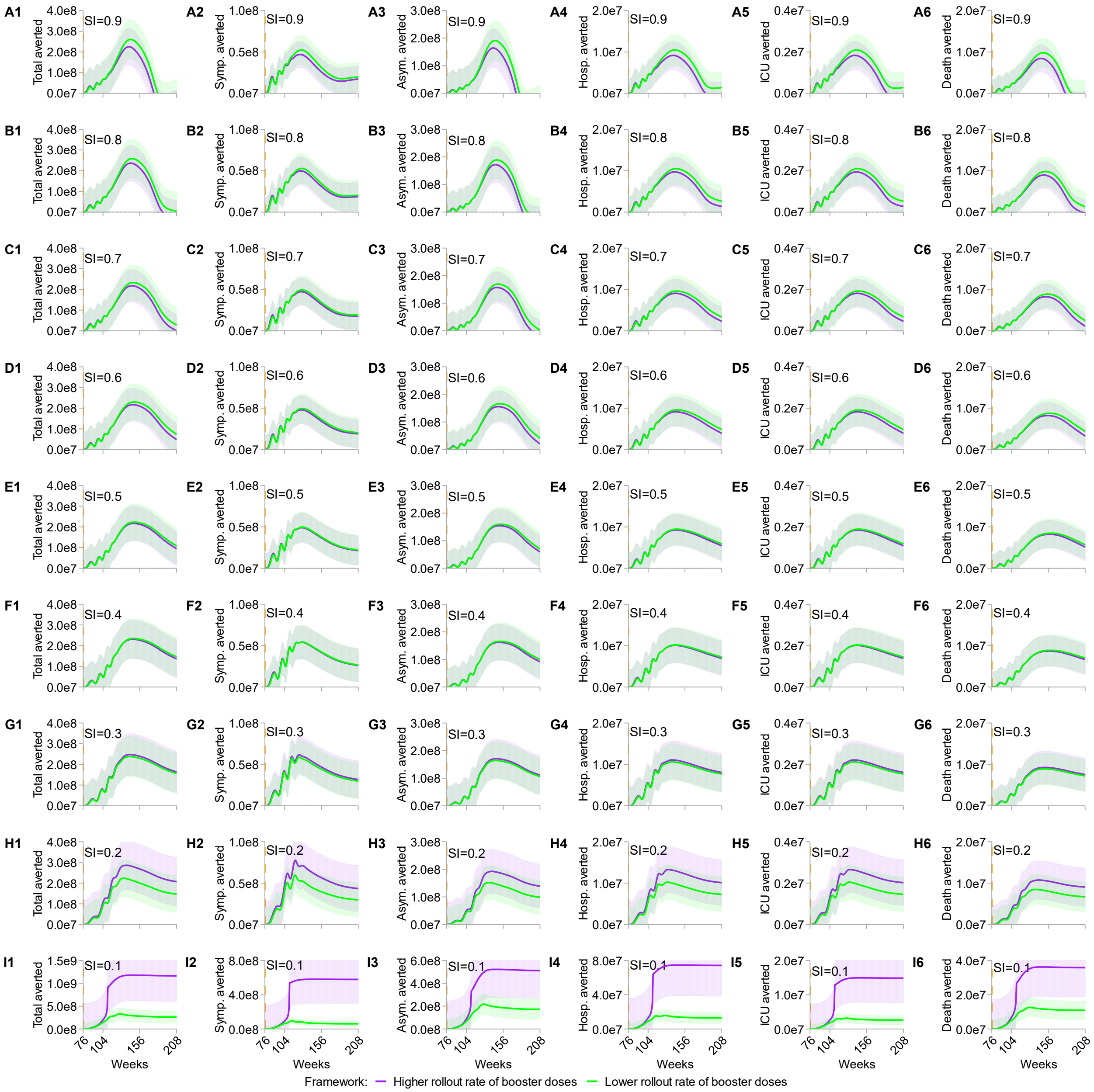


**Figure S16: Sensitivity test on initial infection size (smaller) for six stratified metrics of disease burdens.** SI varies from 0.9 (A1 to A6) to 0.1 (I1 to I6) in 0.1 increments. All scenarios denote the cumulative cases averted over four years and the population size is equal to one billion. "Total averted" represents the total cases averted; "Symp. averted" depicts the symptomatic cases averted; "Asym. averted" represents the asymptomatic cases averted; "Hosp. averted" depicts the hospitalization cases averted; "ICU averted" outlines the averted cases of patients needing intensive care unit; "Death averted" delineates the death cases averted. For each SI scenario, higher (purple curves and shaded areas) and lower (green curves and shaded areas) rollout rates of booster doses are equal to 4 and 0.5 per year respectively. Shaded light colors sketch the 95% confidence intervals of the simulations.

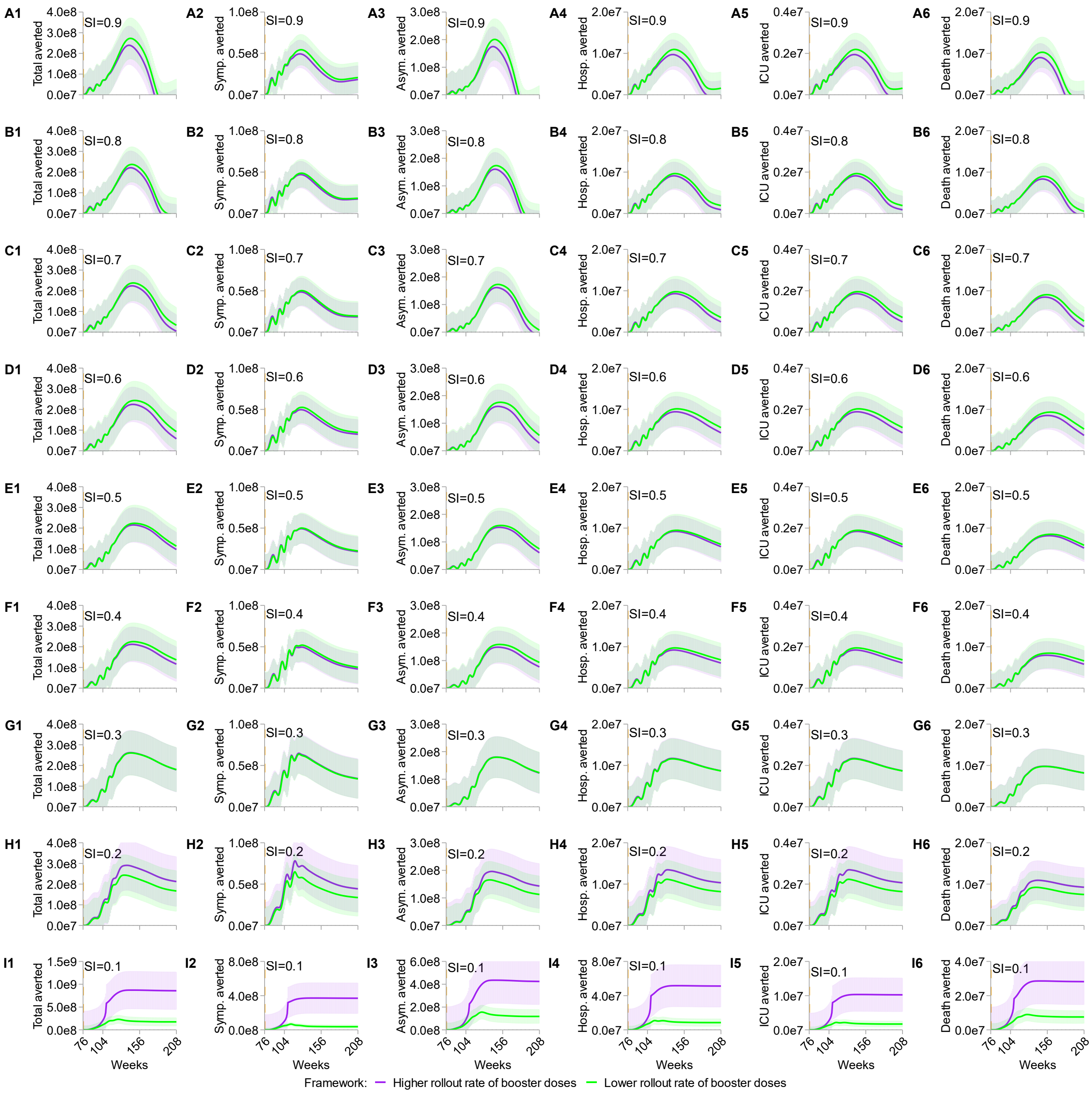


**Figure S17: Sensitivity test on initial infection size (greater) for six stratified metrics of disease burdens.** SI varies from 0.9 (A1 to A6) to 0.1 (I1 to I6) in 0.1 increments. All scenarios denote the cumulative cases averted over four years and the population size is equal to one billion. “Total averted” represents the total cases averted; “Symp. averted” depicts the symptomatic cases averted; “Asym. averted” represents the asymptomatic cases averted; “Hosp. averted” depicts the hospitalization cases averted; “ICU averted” outlines the averted cases of patients needing intensive care unit; “Death averted” delineates the death cases averted. For each SI scenario, higher (purple curves and shaded areas) and lower (green curves and shaded areas) rollout rates of booster doses are equal to 4 and 0.5 per year respectively. Shaded light colors sketch the 95% confidence intervals of the simulations.

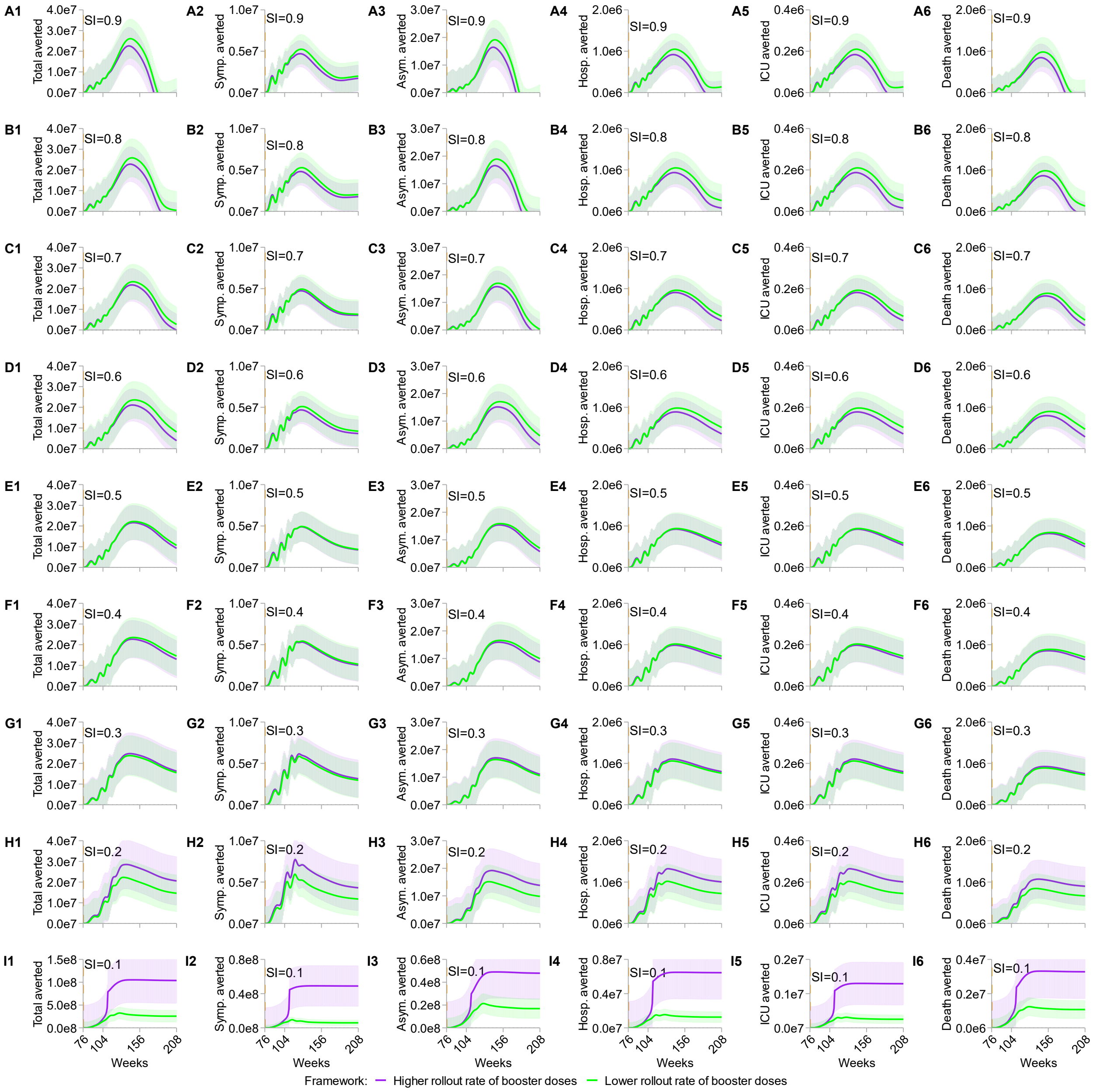


**Figure S18: Sensitivity test on population size (N=1e8) for six stratified metrics of disease burdens.** SI varies from 0.9 (A1 to A6) to 0.1 (I1 to I6) in 0.1 increments. All scenarios denote the cumulative cases averted over four years and the population size is equal to one billion. "Total averted" represents the total cases averted; "Symp. averted" depicts the symptomatic cases averted; "Asym. averted" represents the asymptomatic cases averted; "Hosp. averted" depicts the hospitalization cases averted; "ICU averted" outlines the averted cases of patients needing intensive care unit; "Death averted" delineates the death cases averted. For each SI scenario, higher (purple curves and shaded areas) and lower (green curves and shaded areas) rollout rates of booster doses are equal to 4 and 0.5 per year respectively. Shaded light colors sketch the 95% confidence intervals of the simulations.

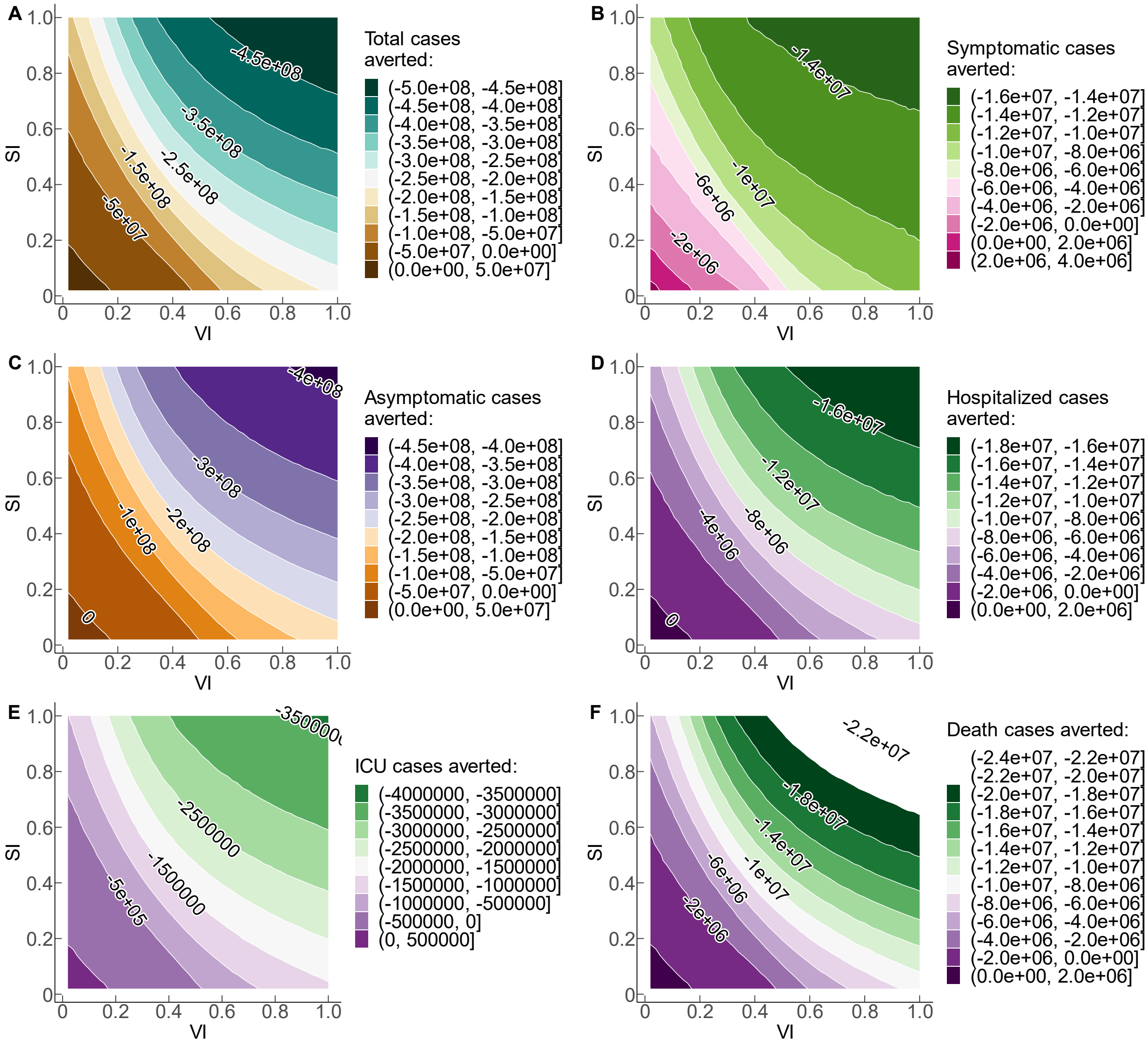


**Figure S19: Trade-off sensitivity test on population size (N=1e8) for six stratified metrics of disease burdens.** SI and VI vary from 0.0 to 1.0 using 50 simulations for each metric. All stratified cases sketch the cumulative cases averted over four years. Positive numbers on the graphs illustrate the positive cases averted, and negative numbers depict the negative trade-off for each health outcome when VS and SI are sub-optimal.

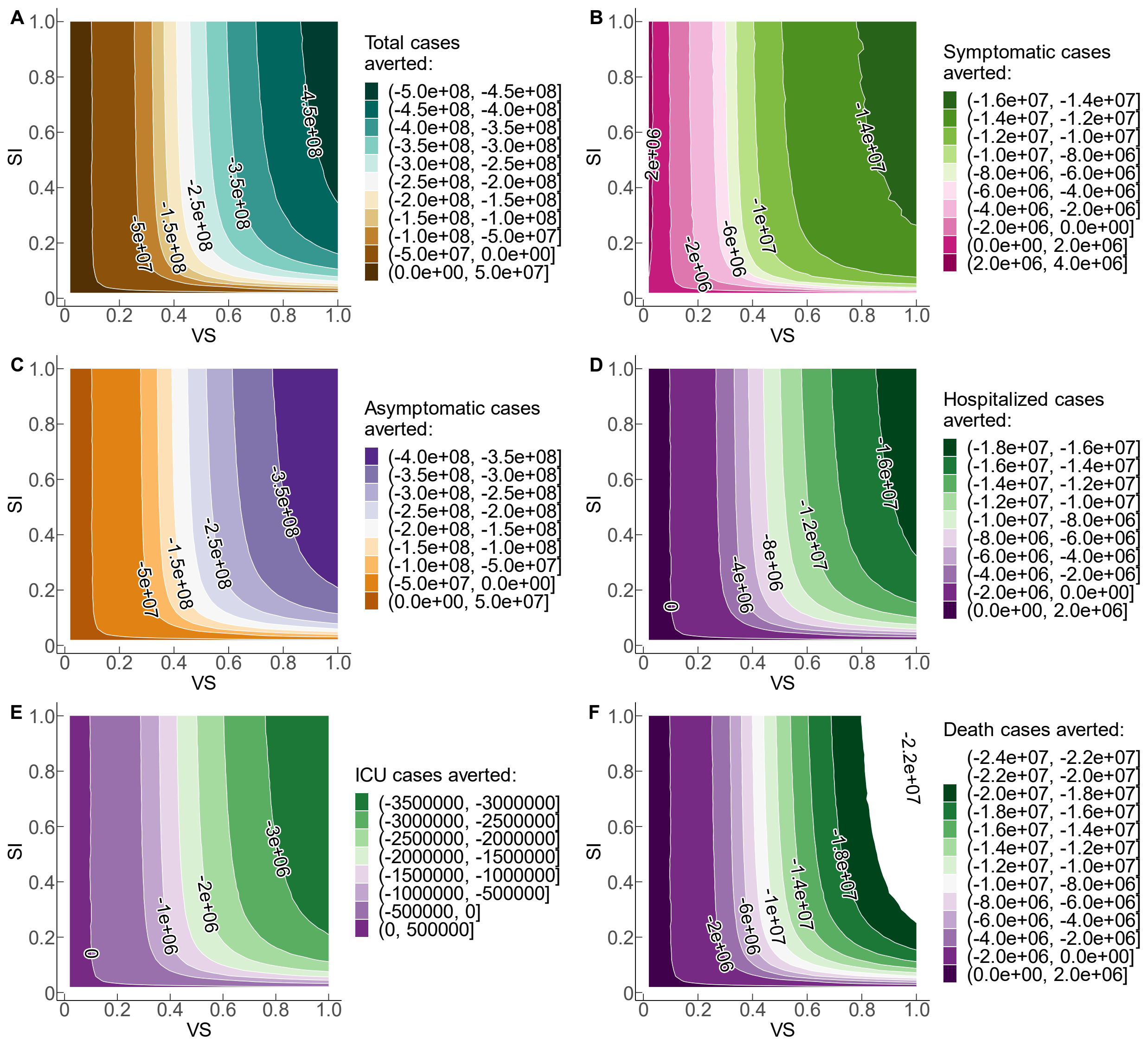


**Figure S20: Trade-off sensitivity test on stratified cases averted with population size equal to 1e8.** Simulation of the cumulative stratified cases averted over four years period for SI and VS ranging from 0 to 1.0 using 50 simulations for each metric respectively. Positive numbers on the graphs illustrate the positive cases averted, and negative numbers depict the negative trade-off for each health outcome when VS and SI are sub-optimal.

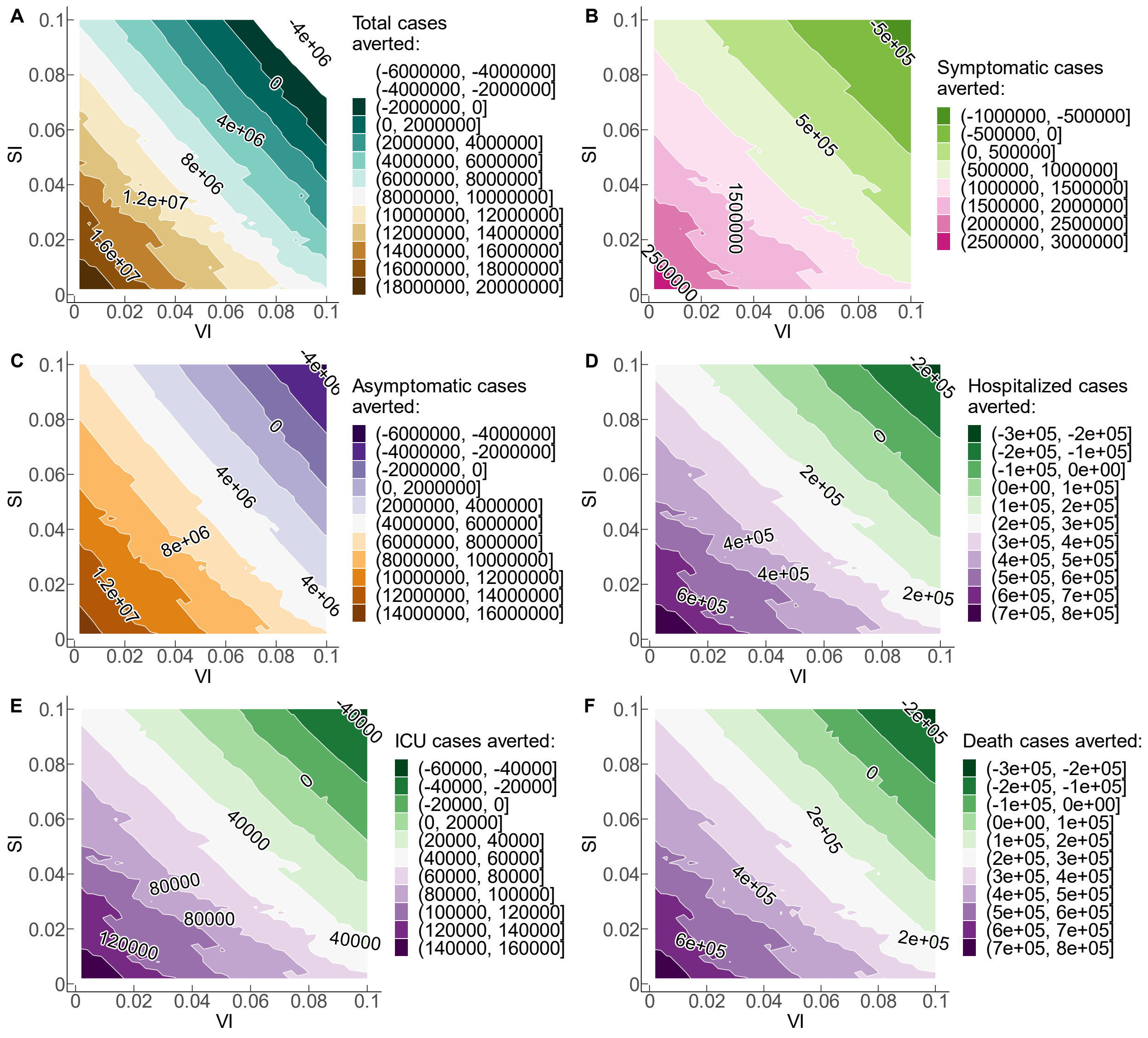


**Figure S21: Trade-off sensitivity test on stratified cases averted with population size equal to 1e8.** Simulation of the stratified cases averted over four years period for SI and VI ranging from 0 to 0.1 using 50 simulations for each metric respectively. Positive numbers illustrate the positive cases averted, and negative numbers depict the negative trade-off for each health burden.

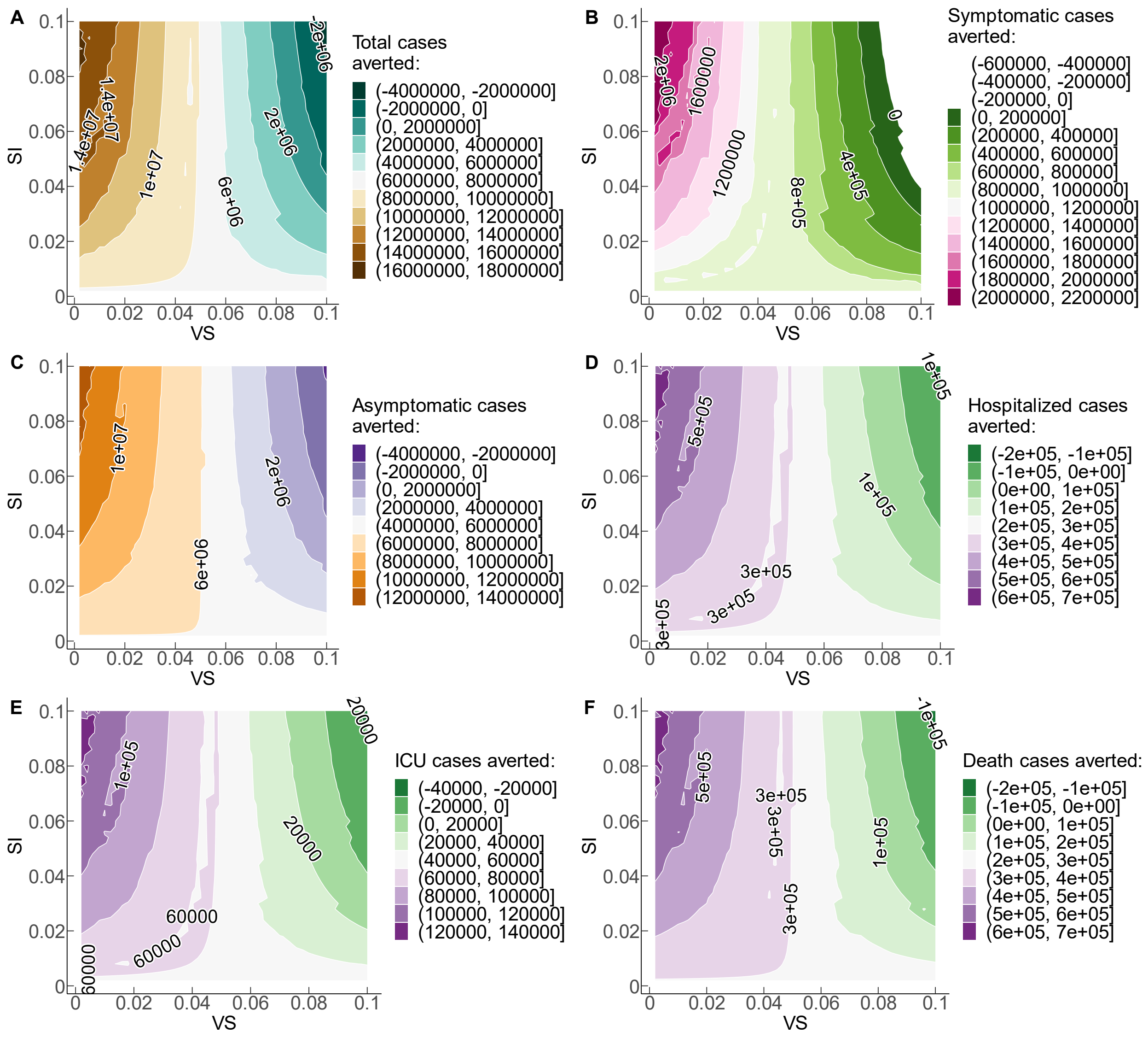


**Figure S22: Trade-off sensitivity test on stratified cases averted with population size equal to 1e8.** Simulation of the stratified cases averted over four years period for SI and VS ranging from 0 to 0.1 using 50 simulations for each metric respectively. Positive numbers illustrate the positive cases averted, and negative numbers depict the negative trade-off for each health burden.

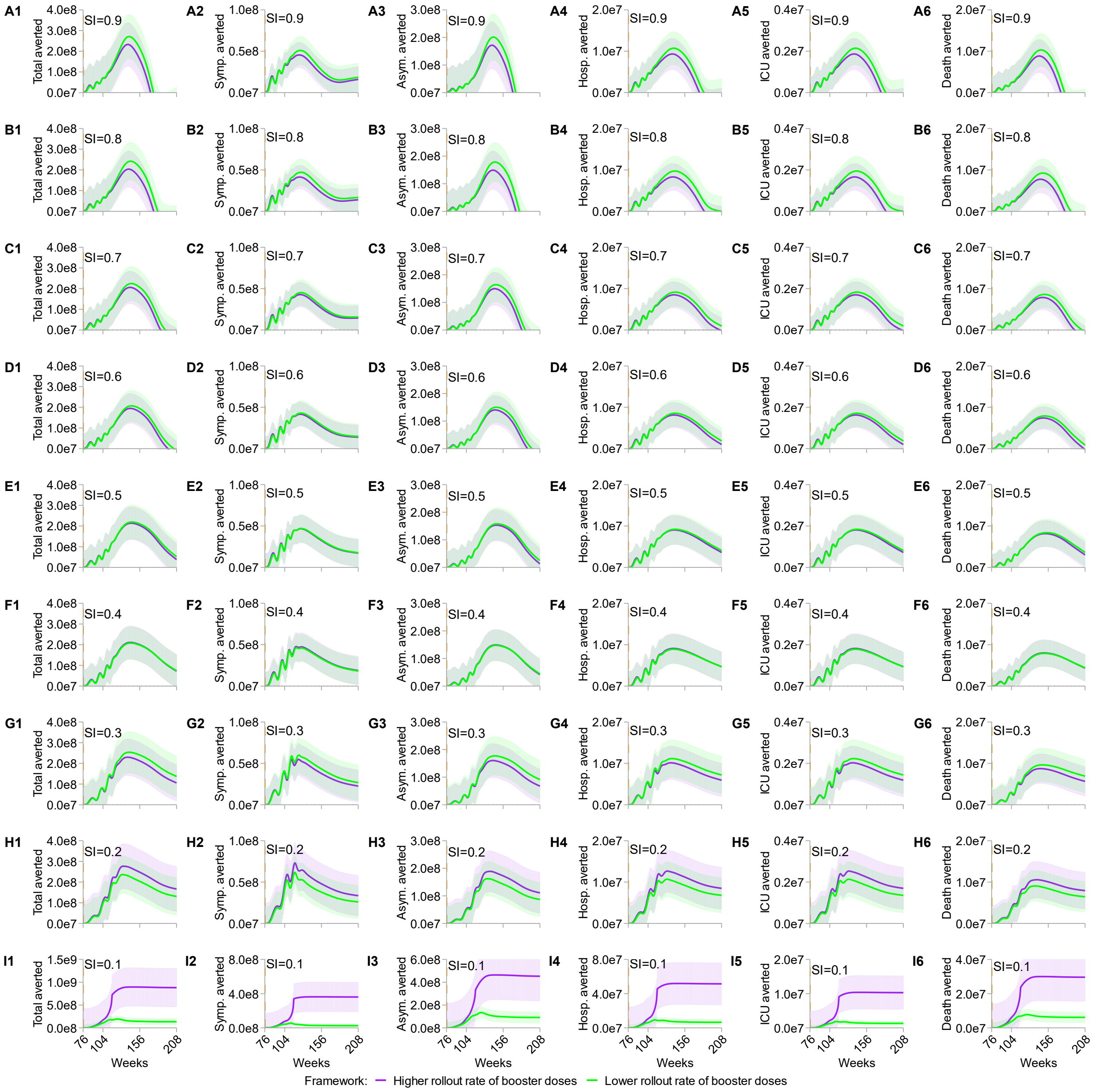


**Figure S23: Multiway sensitivity test on parameters including the rate of infection from susceptibility after immunity of dose 1 wanes, rate of infection from susceptibility after immunity of dose 2 wanes, rate of waning to susceptibility after primary dose 2 vaccination for six stratified metrics of disease burdens.** SI varies from 0.9 (A1 to A6) to 0.1 (I1 to I6) in 0.1 increments. All scenarios denote the cumulative cases averted over four years and the population size is equal to one billion. “Total averted” represents the total cases averted; “Symp. averted” depicts the symptomatic cases averted; “Asym. averted” represents the asymptomatic cases averted; “Hosp. averted” depicts the hospitalization cases averted; “ICU averted” outlines the averted cases of patients needing intensive care unit; “Death averted” delineates the death cases averted. For each SI scenario, higher (purple curves and shaded areas) and lower (green curves and shaded areas) rollout rates of booster doses are equal to 4 and 0.5 per year respectively. Shaded light colors sketch the 95% confidence intervals of the simulations.

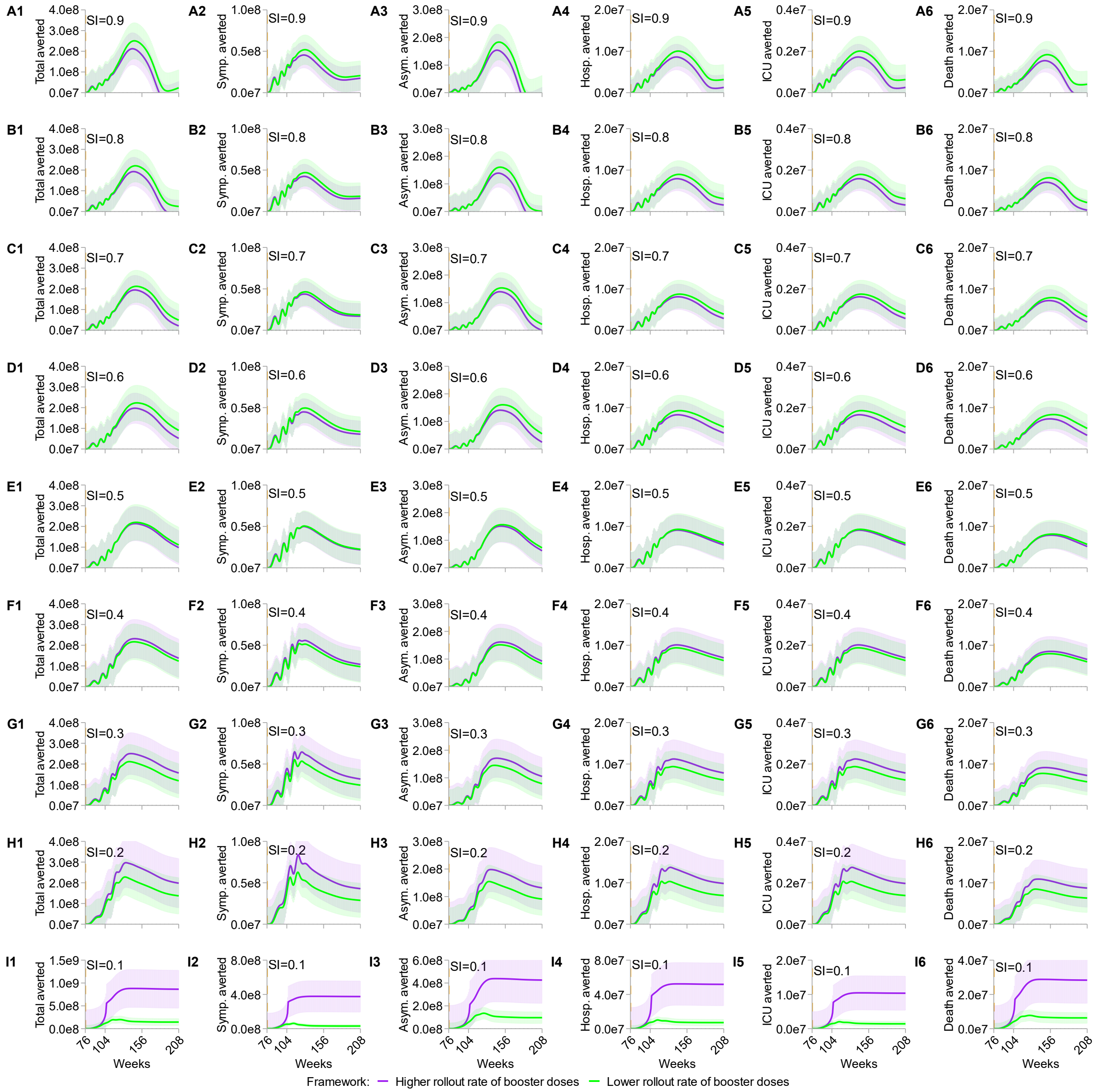


**Figure S24: Multiway sensitivity test on parameters including the rate of infection after dose 1 vaccination, rate of infection after dose 2 vaccination, rate of waning to susceptibility after primary dose 1 vaccination, rate of waning to secondary susceptibility after recovery for six stratified metrics of disease burdens.** SI varies from 0.9 (A1 to A6) to 0.1 (I1 to I6) in 0.1 increments. All scenarios denote the cumulative cases averted over four years and the population size is equal to one billion. “Total averted” represents the total cases averted; “Symp. averted” depicts the symptomatic cases averted; “Asym. averted” represents the asymptomatic cases averted; “Hosp. averted” depicts the hospitalization cases averted; “ICU averted” outlines the averted cases of patients needing intensive care unit; “Death averted” delineates the death cases averted. For each SI scenario, higher (purple curves and shaded areas) and lower (green curves and shaded areas) rollout rates of booster doses are equal to 4 and 0.5 per year respectively. Shaded light colors sketch the 95% confidence intervals of the simulations.

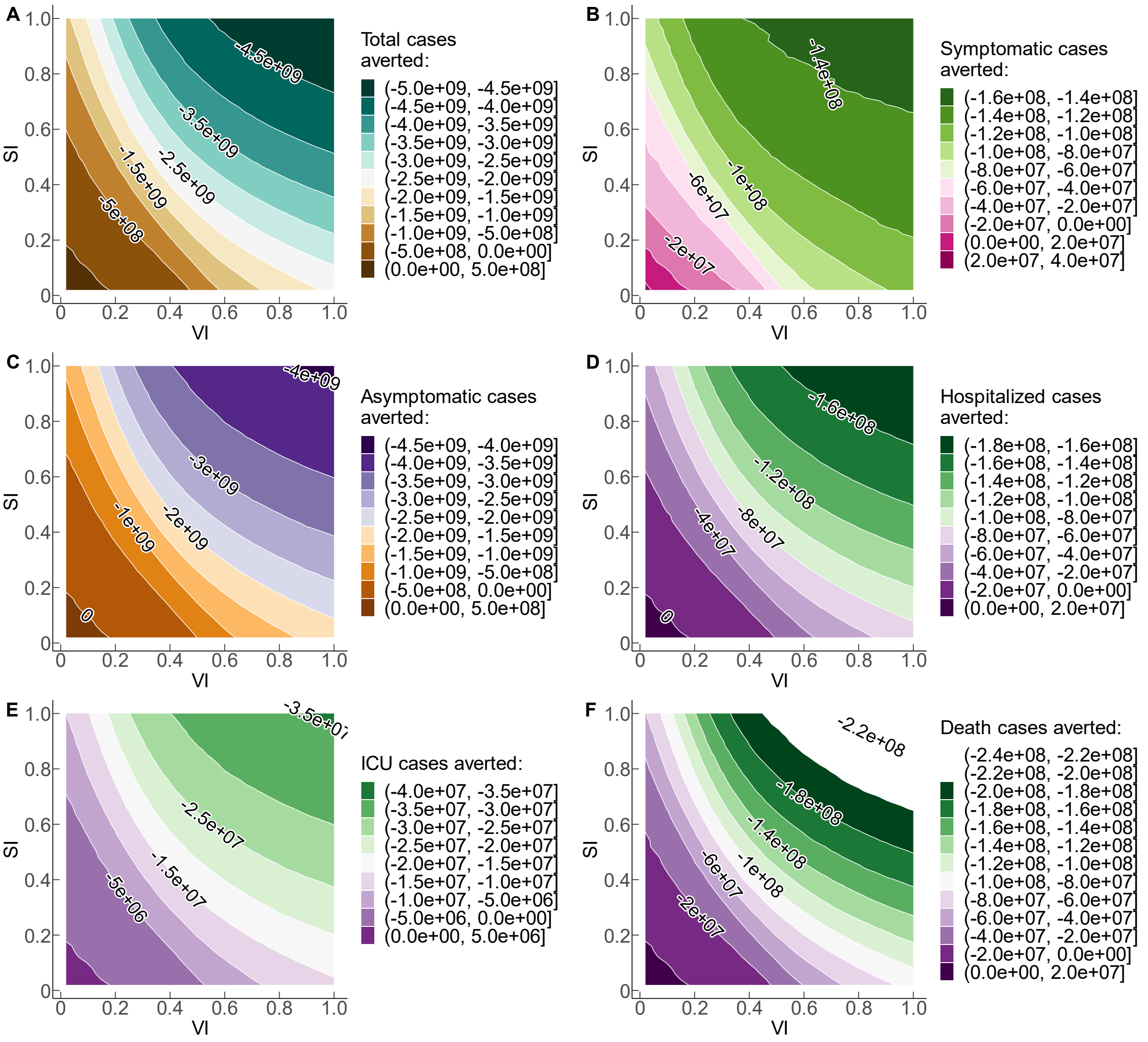


**Figure S25: Simulation trade-offs of disease burdens varying SI and VI from 0 to 1.** Simulation of the cumulative stratified cases averted over four years with SI and VI ranging from 0 to 1.0 using 50 simulations for each metric respectively. The population size is equal to 1e9. Positive numbers on each curve illustrate the positive cases averted for each health outcome, and negative numbers depict the negative trade-off when VI and SI transform to sub-optimal ceteris paribus. The rollout rate of booster doses is 0.5 per year, representing the non-productive rollout scenario.

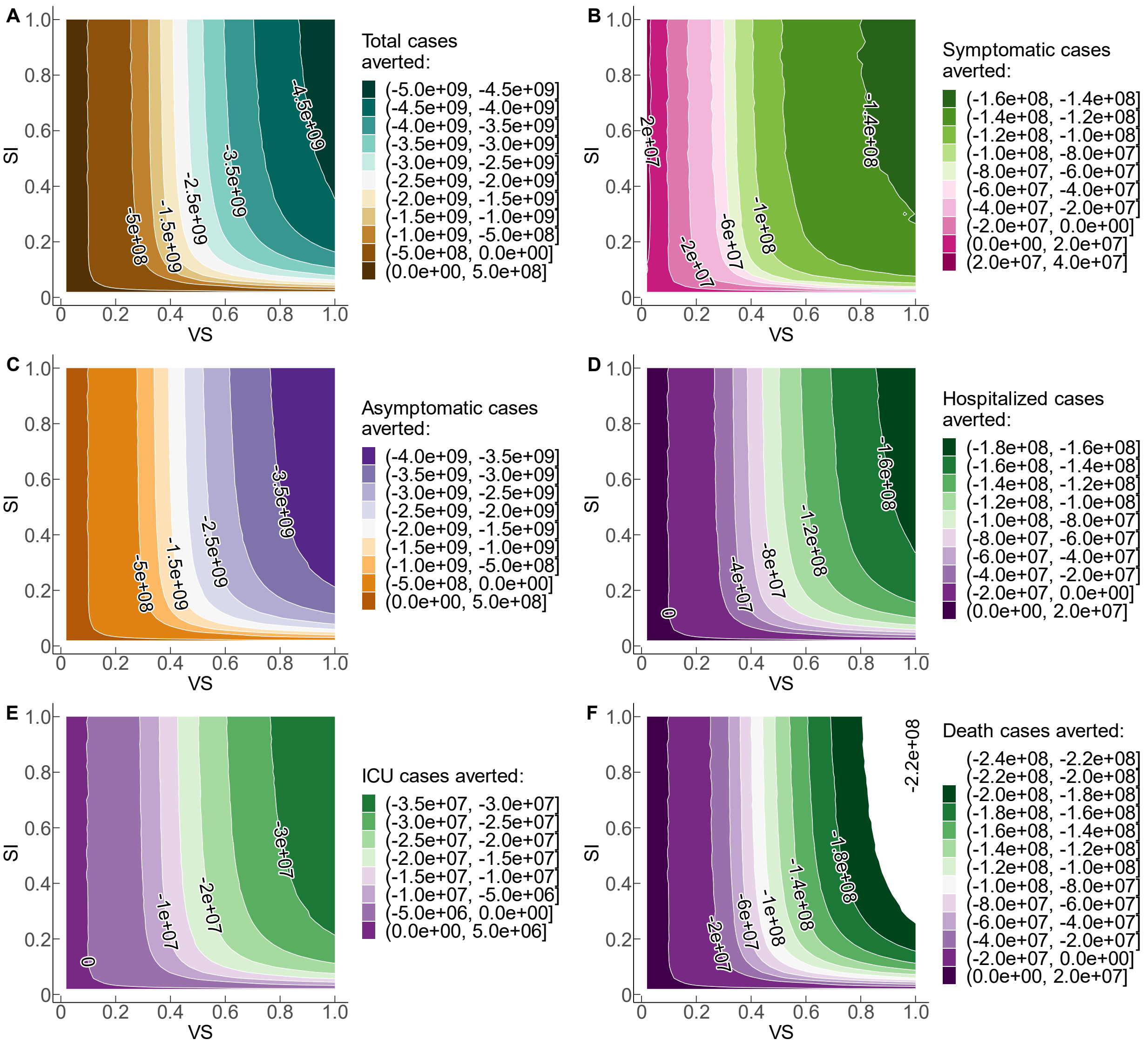


**Figure S26: Simulation trade-offs of disease burdens with varying immunity profiles using VS and SI.** Simulation of the cumulative stratified cases averted over four years with SI and VS ranging from 0 to 1.0 using 50 simulations for each metric respectively. The population size is equal to 1e9. Positive numbers on each curve illustrate the positive cases averted for each health burden, and negative numbers depict the negative trade-off when VI and SI are sub-optimal ceteris paribus. The rollout rate of booster doses is 0.5 per year, representing the non-productive rollout scenario.

**Table S1.**

**Table S1. Key simulation parameters for Fig. 3**

| Name | Description | Baseline value | References |
|---|---|---|---|
| $\boldsymbol{N}$ | population size | 1e9 | Assumed |
| $\alpha$ | relative infectiousness of $I_S$ versus $I_P$ | 1 | Assumed |
| $\beta$ | rate of transmission | $R*\gamma$ | [20-23] |
| $\alpha_V$ | relative infectiousness of $I_V$ versus $I_P$ | 1 | Assumed |
| $\alpha_1$ | relative infectiousness of $I_{S_1}$ versus $I_P$ | 1 | Assumed |
| $\alpha_2$ | relative infectiousness of $I_{S_2}$ versus $I_P$ | 1 | Assumed |
| $\alpha_3$ | relative infectiousness of $I_{S_3}$ versus $I_P$ | 1 | Assumed |
| $\varepsilon_1$ | rate to infection from susceptibility after immunity of dose 1 wanes | 0.5 per year | [20-23] |
| $\varepsilon_2$ | rate to infection from susceptibility after immunity of dose 2 wanes | 0.3 per year | [20-23] |
| $\varepsilon_3$ | rate to infection from susceptibility after immunity of dose 3 wanes | 0.1 per year | Assumed |
| $\varepsilon_{V_1}$ | rate of infection after dose 1 vaccination | 0.2 per year | [20-23] |
| $\varepsilon_{V_2}$ | rate of infection after dose 2 vaccination | 0.05 per year | [20-23] |
| $\varepsilon_{V_3}$ | rate of infection after dose 3 vaccination | 0.05 per year | Assumed |
| $\boldsymbol{\rho_1}$ | rate of waning to susceptibility after primary dose 1 vaccination | 8 per week. | [20-23] |
| $\boldsymbol{\rho_2}$ | rate of waning to susceptibility after primary dose 2 vaccination | 2/3 per week | [20-23] |
| $\boldsymbol{\rho_3}$ | rate of waning to susceptibility after booster dose 3 vaccination | 2 per week | Assumed |
| $\mu$ | birth rate or death rate of population | 50/1000 per year | [20-23] |
| $\gamma$ | recovery rate of infected patients | 1.4 per year | [20-23] |
| $\boldsymbol{\nu}$ | vaccination rate of primary dose 1 | 0.01 per year | [20-23] |
| $\omega$ | vaccination rate of primary dose 2 | 0.05 per year | [20-23] |
| $\omega_1$ | vaccination rate of booster dose 3 | {0.5,4} per year | Assumed |
| $\delta$ | rate of waning to secondary susceptibility after recovery | 1/4 per year. | [20-23] |
| $S_{vax_{1-3}}$ | initiation timing of dose 1&2&3 | {44,52,76} [weeks] | Assumed |
| $\boldsymbol{S_{P0}}$ | initial size of the full susceptible population | $1 - I_{S0} - \boldsymbol{I_{P0}}$ | [20-22,26-28] |
| $\boldsymbol{I_{S0}}$ | initial ratio of secondary infections relative to primary infections | 0 | [20-26] |
| $\boldsymbol{I_{P0}}$ | initial size of primary infections | 1e-9 | [20-23] |
| $R$ | reproduction number | 2.3[seasonal variation] | [20-22,25-29] |
| $c,d$ | partially susceptible individuals vaccinated | 0.01 | [20-23] |

**Table S2.**

**Table S2. Kay simulation parameters for Fig. 4**

| **Name** | Description | Baseline value | References |
|---|---|---|---|
| $\boldsymbol{N}$ | population size | 1e9 | Assumed |
| $\alpha$ | relative infectiousness of $I_S$ versus $I_P$ | 1 | Assumed |
| $\beta$ | rate of transmission | $R*\gamma$ | [20-23] |
| $\alpha_V$ | relative infectiousness of $I_V$ versus $I_P$ | 1 | Assumed |
| $\alpha_1$ | relative infectiousness of $I_{S_1}$ versus $I_P$ | 1 | Assumed |
| $\alpha_2$ | relative infectiousness of $I_{S_2}$ versus $I_P$ | 1 | Assumed |
| $\alpha_3$ | relative infectiousness of $I_{S_3}$ versus $I_P$ | 1 | Assumed |
| $\varepsilon_1$ | rate to infection from susceptibility after immunity of dose 1 wanes | 0.5 per year | [20-23] |
| $\varepsilon_2$ | rate to infection from susceptibility after immunity of dose 2 wanes | 0.3 per year | [20-23] |
| $\varepsilon_3$ | rate to infection from susceptibility after immunity of dose 3 wanes | [0,1] in 50 simulations | Assumed |
| $\varepsilon_{V_1}$ | rate of infection after dose 1 vaccination | 0.2 per year | [20-23] |
| $\varepsilon_{V_2}$ | rate of infection after dose 2 vaccination | 0.05 per year | [20-23] |
| $\varepsilon_{V_3}$ | rate of infection after dose 3 vaccination | [0,1] in 50 simulations | Assumed |
| $\boldsymbol{\rho_1}$ | rate of waning to susceptibility after primary dose 1 vaccination | 8 per week. | [20-23] |
| $\boldsymbol{\rho_2}$ | rate of waning to susceptibility after primary dose 2 vaccination | 2/3 per week | [20-23] |
| $\boldsymbol{\rho_3}$ | rate of waning to susceptibility after booster dose 3 vaccination | 2 per week | Assumed |
| $\mu$ | birth rate or death rate of population | 50/1000 per year | [20-23] |
| $\gamma$ | recovery rate of infected patients | 1.4 per year | [20-23] |
| $\boldsymbol{\nu}$ | vaccination rate of primary dose 1 | 0.01 per year | [20-23] |
| $\omega$ | vaccination rate of primary dose 2 | 0.05 per year | [20-23] |
| $\omega_1$ | vaccination rate of booster dose 3 | {0.5,4} per year | Assumed |
| $\delta$ | rate of waning to secondary susceptibility after recovery | 1/4 per year. | [20-23] |
| $S_{vax_{1-3}}$ | initiation timing of dose 1&2&3 | {44,52,76} [weeks] | Assumed |
| $\boldsymbol{S_{P0}}$ | initial size of the full susceptible population | $1 - I_{S0} - \boldsymbol{I_{P0}}$ | [20-22,26-28] |
| $\boldsymbol{I_{S0}}$ | initial ratio of secondary infections relative to primary infections | 0 | [20-26] |
| $\boldsymbol{I_{P0}}$ | initial size of primary infections | 1e-9 | [20-23] |
| $R$ | reproduction number | 2.3[seasonal variation] | [20-22,25-29] |
| $c,d$ | partially susceptible individuals vaccinated | 0.01 | [20-23] |

**Table S3.**

**Table S3. Simulation parameters for Fig. 5**

| **Name** | Description | Baseline value | References |
|---|---|---|---|
| $\boldsymbol{N}$ | population size | 1e9 | Assumed |
| $\alpha$ | relative infectiousness of $I_S$ versus $I_P$ | 1 | Assumed |
| $\beta$ | rate of transmission | $R*\gamma$ | [20-23] |
| $\alpha_V$ | relative infectiousness of $I_V$ versus $I_P$ | 1 | Assumed |
| $\alpha_1$ | relative infectiousness of $I_{S_1}$ versus $I_P$ | 1 | Assumed |
| $\alpha_2$ | relative infectiousness of $I_{S_2}$ versus $I_P$ | 1 | Assumed |
| $\alpha_3$ | relative infectiousness of $I_{S_3}$ versus $I_P$ | 1 | Assumed |
| $\varepsilon_1$ | rate to infection from susceptibility after immunity of dose 1 wanes | 0.5 per year | [20-23] |
| $\varepsilon_2$ | rate to infection from susceptibility after immunity of dose 2 wanes | 0.3 per year | [20-23] |
| $\varepsilon_3$ | rate to infection from susceptibility after immunity of dose 3 wanes | [0, 1] in 50 simulations | Assumed |
| $\varepsilon_{V_1}$ | rate of infection after dose 1 vaccination | 0.2 per year | [20-23] |
| $\varepsilon_{V_2}$ | rate of infection after dose 2 vaccination | 0.05 per year | [20-23] |
| $\varepsilon_{V_3}$ | rate of infection after dose 3 vaccination | 0.05 per year | Assumed |
| $\boldsymbol{\rho_1}$ | rate of waning to susceptibility after primary dose 1 vaccination | 8 per week. | [20-23] |
| $\boldsymbol{\rho_2}$ | rate of waning to susceptibility after primary dose 2 vaccination | 2/3 per week | [20-23] |
| $\boldsymbol{\rho_3}$ | rate of waning to susceptibility after booster dose 3 vaccination | [0, 1] in 50 simulations | Assumed |
| $\mu$ | birth rate or death rate of population | 50/1000 per year | [20-23] |
| $\gamma$ | recovery rate of infected patients | 1.4 per year | [20-23] |
| $\boldsymbol{\nu}$ | vaccination rate of primary dose 1 | 0.01 per year | [20-23] |
| $\omega$ | vaccination rate of primary dose 2 | 0.05 per year | [20-23] |
| $\omega_1$ | vaccination rate of booster dose 3 | {0.5,4} per year | Assumed |
| $\delta$ | rate of waning to secondary susceptibility after recovery | 1/4 per year. | [20-23] |
| $S_{vax_{1-3}}$ | initiation timing of dose 1&2&3 | {44,52,76} [weeks] | Assumed |
| $\boldsymbol{S_{P0}}$ | initial size of the full susceptible population | $1 - I_{S0} - \boldsymbol{I_{P0}}$ | [20-22,26-28] |
| $\boldsymbol{I_{S0}}$ | initial ratio of secondary infections relative to primary infections | 0 | [20-26] |
| $\boldsymbol{I_{P0}}$ | initial size of primary infections | 1e-9 | [20-23] |
| $R$ | reproduction number | 2.3[seasonal variation] | [20-22,25-29] |
| $c,d$ | partially susceptible individuals vaccinated | 0.01 | [20-23] |

**Data S1. (separate file)**
Supplementary Data file S1.xlsx.
